# Mpox clinical and epidemiological patterns in the Central African Republic: a systematic review and meta-analysis

**DOI:** 10.1101/2025.07.09.25331215

**Authors:** Fabrice Zobel Lekeumo Cheuyem, Davy Roméo Takpando-le-grand, Chabeja Achangwa, Lionel Berthold Keubou Boukeng, Solange Dabou, Henri Donald Mutarambirwa, Raissa Katy Noa Otsali, Ariane Nouko, Michel Franck Edzamba, Martine Golda Mekouzou Tsafack, Armel Evouna Mbarga, Georges Nguefack-Tsague

## Abstract

**Background:** Mpox remains a critical public health challenge in Central Africa, where endemic circulation of the virulent Clade I virus persists through zoonotic and human-to-human transmission. This systematic review and meta-analysis synthesize 40 years of evidence (1984-2024) to characterize Mpox epidemiology, vaccination gaps and clinical pattern in the Central African Republic (CAR), informing targeted control strategies for this re-emerging threat.

**Methods:** Following PRISMA guidelines, we conducted a comprehensive search of PubMed, Scopus, ScienceDirect, Web of science, Embase, Cochrane Library, and AJOL for studies reporting laboratory-confirmed Mpox cases in CAR. Pooled estimates were calculated using random/fixed-effects models with R Software version 4.4.2 with subgroup analysis conducted for period, region, participants and disease burden. Heterogeneity (*I*^2^) and publication bias (funnel plots, Egger’s and Begg’s tests) were rigorously assessed. The confidence intervals (CIs) were estimated at 95% level of confidence. A *p*-value ⍰5% was considered statistically significant.

**Results:** Analysis of seven studies (1984-2023) found a pooled severity rate of 60.9% (95%CI: 48.5-77.7), peaking at 77.3% (95%CI: 55.6-90.2) in Health Region 6. Before the global mpox outbreak, the CFR for confirmed cases was 11.54% (95% CI: 6.11-20.71). Eastern Health Regions had a higher CFR of 11.11% (95% CI: 5.08-22.60), while Western Regions reported 0.00% (95% CI: 0.00-100.00). For suspected cases, the CFR was 12.77% (95% CI: 7.39-21.15), slightly declining to 9.09% (95% CI: 0.23-41.28) post-2022. This geographic disparity remained, with Eastern Regions at 10.17% (95% CI: 4.64-20.84) and Western Regions at 5.26% (95% CI: 0.74-29.39). Vaccination uptake in CAR was only 20.00% (95% CI: 10.33-35.17). The clinical profile included fever (91.1% (95%CI: 42.9-99.3)), rash (85.5% (95%CI: 75.7-91.8)), and lymphadenopathy (57.0% (95%CI: 34.3-77.1)).

**Conclusion:** The CAR faces a disproportionate Mpox burden characterized by high severity, elevated mortality, and suboptimal vaccine coverage, particularly in eastern regions. These findings suggest the urgent need for: (1) enhanced surveillance systems with genomic sequencing capacity; (2) prevention of disease lethality through early case detection and appropriate care provision; (3) strengthening vaccine distribution prioritizing high-risk populations; and (4) context-specific interventions, including community education and healthcare worker training, to curb future outbreaks.

## Background

Mpox, formerly known as monkeypox, is an infectious viral disease caused by the monkeypox virus [1]. In 2022, the World Health Organization (WHO) declared Mpox a Public Health Emergency of International Concern for the second time, confirming its continued threat to global health security and underscoring the persistent risks posed by the disease [2]. As of May 2025, a total of 133 countries globally have notified 142,151 confirmed Mpox cases with 328 deaths, yielding a case fatality rate (CFR) of 0.09% [3].

By June 2025, several African countries have reported community transmission of mpox due to clade Ib monkeypox virus in the past six weeks [4]. These include Burundi, the Democratic Republic of the Congo, Ethiopia, Kenya, Malawi, Rwanda, South Sudan, Tanzania, Uganda, and Zambia, all of which are experiencing ongoing human-to-human transmission of the virus. The Democratic Republic of the Congo continues to report the highest number of confirmed mpox cases in Africa in 2025 [5]. Mpox was first identified in the DRC, but since then, it has spread to nearby non-endemic African nations [6]. Such countries include the Central African Republic (CAR) and the Republic of the Congo, where investigations have provided evidence of sustained community transmission [7,8].

To adequately respond to this community transmission, the CAR health authorities’ response to this public health threat should rely on solid surveillance and healthcare systems prepared to detect, diagnose, and promptly manage all notified cases to avoid preventable deaths in-hospital or within the community [9]. This is a significant challenge, as the complexity and severity of disease outbreaks are placing a heavy burden on under-resourced health systems like CAR’s. In addition, marginalized and hard-to-reach populations, such as refugees, internally displaced persons, nomadic population and migrants, frequently encounter significant obstacles in accessing healthcare. This lack of access increases their susceptibility to this reemerging disease [10].

The Central African Republic’s (CAR) situation regarding this outbreak is particularly sensitive due to its socioeconomic and political condition. The country is one of the least developed globally, with decades of political instability, armed conflict, and widespread displacement severely impacting its socio-economic status [11]. This has led to high levels of poverty, food insecurity, and a significant proportion of the population living in precarious conditions, often with limited access to basic amenities like clean water and sanitation [12]. These socio-economic vulnerabilities can exacerbate disease transmission and hinder effective public health responses, as communities often rely on traditional healers and informal healthcare providers. This reliance can delay diagnosis and appropriate management of infectious diseases like Mpox [13,14]. In this case, prevention remains one of the key strategies to control the spread of the disease within hotspots and across the country.

The implementation of Mpox vaccination activities could be an effective way to respond to this outbreak by vaccinating populations most at risk of contracting Mpox [15,16]. This includes individuals living in close proximity to wildlife reservoirs, particularly in forested, rural, and remote areas that experience a severe shortage of qualified health personnel, medical supplies, and diagnostic capabilities [8].

Most previous studies on Mpox in CAR were conducted with small sample sizes, thereby limiting the generalizability of their findings across the country [8,17–19]. Understanding the evolution of this disease and its regional distribution in this intricate setting is essential for designing and implementing effective Mpox prevention and control strategies tailored to the realities of the CAR. Therefore, this study aimed to provide insight into the epidemiological and clinical profile of the multiple Mpox outbreaks reported in CAR.

## Methods

### Study design

This is the first systematic review and meta-analysis providing insight on the clinical and epidemiological profile of Mpox in CAR from 1984 to 2024. It was reported based on the Preferred Reporting Items for Systematic Review and Meta-analysis (PRISMA) guidelines [20].

### Article searching strategy

Literature searches were performed on PubMed, Scopus, ScienceDirect, Web of Science, Embase, Cochrane Library, and AJOL to identify published research. The search approach involved the title and abstract screening of each study. A synthesis of keywords and Medical Subject Headings (MeSH) vocabulary was utilized, employing Boolean logic operators to narrow the search for each database used (Additional File 1, Supplementary Table 2). To ensure thoroughness, a supplementary manual searching was conducted to identify additional relevant publications not found in electronic databases such as Google scholar (first 1000 entries assessed and no filter applied). Additionally, the reference lists of included studies were reviewed to identify further relevant articles. Last literature search was concluded on February, 2025.

### Study setting

The CAR is a landlocked nation situated in the heart of Africa, with an estimated population of 6 million inhabitants in 2024 [21]. Most of its population (62%) lives in rural areas, with women being the most represented (50.9%). The country covers an area of 623,000 km^2^ and is bordered to the North by Chad, to the East by South Sudan and the Republic of Sudan, to the South by the Democratic Republic of Congo and the Republic of Congo, and to the West by Cameroon [22]. Administratively, the country is divided into seven Health Regions, 20 Divisions, 85 Sub-divisions, 177 Councils, and around 8,302 villages. The capital, Bangui, has 10 Sub-divisions [23]. The CAR is characterized by diverse geographical features, ranging from dense rainforests in the south to savanna and semi-arid zones in the north [24]. This varied landscape, coupled with a predominantly rural population, influences human-animal interactions.

### Eligibility criteria

#### Inclusion criteria

This systematic review and meta-analysis encompassed all published reports, regardless of study type or design, which documented Mpox in CAR. Only articles published in English or French were included, and no temporal restrictions were applied to the publication date.

#### Exclusion criteria

Studies were deemed ineligible for inclusion if their research focus diverged from our investigative objectives or overlap with other existing results. In addition, letters to editor, commentary or reports not specifying the sample size were not included.

### Data extraction

A structured Microsoft Office Excel 2016 form was used to collect data from all included articles. The data extraction checklist included the first author’s name, study year, region, study design, setting, number of suspected, confirmed, severe, and death cases, the number of individuals who received the Mpox vaccine, sample size, and the number of events of each clinical manifestation. Two authors independently assessed each article for quality and relevance. Discrepancies between reviewers were resolved through discussion with a third reviewer to achieve consensus.

### Data quality assessment

The quality of included studies was assessed using the Joanna Briggs Institute quality assessment tool for cross-sectional studies generated following outbreak surveillance and investigations activities [25]. For cross-sectional studies, assessment criteria encompassed studies clear definition of inclusion criteria, comprehensive descriptions of study subjects and settings, validity and reliability of exposure measurements, use of objective, standardized criteria for outcome assessment, identification potential confounding factors, implementation of appropriate strategies to address them, and the appropriateness of statistical methods. A binary score of 0 (no or unclear) or 1 (yes) was given after evaluation of each criterion. The overall risk of bias was categorized as low (>50%), moderate (>25-50%), or high (≤25%).

### Outcome measurement

The primary outcomes of this systematic review and meta-analysis were Mpox severity and CFR. The secondary outcomes included the vaccination uptake and the clinical manifestations among suspected and confirmed cases. Mpox severity rate was determined by calculating the proportion of participants exhibiting severe or grave clinical manifestations of Mpox out of the total number of confirmed cases. CFR was calculated by dividing the number of deaths observed within the population of suspected or confirmed cases. The Mpox vaccination uptake rate/coverage was obtained by dividing the total number of people who received the vaccine by the number of eligible participants. The frequency of each clinical manifestation was computed by dividing the number of events of each reported symptom or observed sign by the number of suspected or confirmed cases.

### Operational definition

In primary studies included, a suspected monkeypox case was defined as an individual presenting with a vesicular or pustular rash characterized by deep-seated, firm pustules, and at least one of the following symptoms: fever preceding the eruption, lymphadenopathy (inguinal, axillary, or cervical), or pustules or crusts on the palms of the hands or soles of the feet. A case was defined as laboratory-confirmed Mpox if at least one specimen yielded a positive result in the *Orthopoxvirus*-specific assay, Mpox-specific real-time polymerase chained reaction, or Mpox in culture [26,27]. Cases were classified as severe or grave (requiring hospitalization) if one or more of the following were present: extensive lesions (>100), hemorrhagic or pustular lesions, mucosal involvement (oral, genital, conjunctival), or systemic complications. Systemic complications included high fever (>39°C), altered general state (bedridden), sepsis, encephalitis, secondary bacterial infections, hypotension, septic shock, severe dehydration, and keratitis potentially leading to corneal scarring and blindness [5,26,28]. Vaccination uptake refers to individuals who declared having received an Mpox vaccine or presented an Mpox vaccination card or a scar characteristic of smallpox vaccine injection [15,29].

### Statistical analysis and synthesis

The *I*^2^ statistic was used to assess heterogeneity of pooled estimates between studies. It was categorized as low (<25%), moderate (25−75%), or high (>75%). Subgroup analyses were conducted for temporal trends (study period), geographical disparities (study location), and disease burden. The disease burden cutoff was set based on the median of suspected or confirmed cases reported across included studies. A fixed-effect model was used when heterogeneity was <50%. Timeframes for this analysis were defined based on the global outbreak that led to a heightened global response [30]. Generalized Linear Mixed Models (GLMM), coupled with the Probit-Logit Transformation (PLOGIT), were employed. This approach is ideal for meta-analyses of binary or proportion data because it directly models binomial outcomes and inherently handles studies with 0% or 100% events without needing continuity corrections. A *p*-value of <0.05 was considered significant. All analyses were performed using the ‘meta’ package in R Software version 4.4.2 [31].

### Publication bias and sensitivity test

A visual analysis using funnel plot was carried out to assess publication bias. Absence of publication bias was assumed based on observed symmetry of the inverted funnel shape. To further investigate potential bias, Egger’s regression and Begg’s rank correlation tests were performed. Sensitivity analysis was conducted by iteratively excluding one study at a time to assess the robustness of the pooled estimate.

## Results

From an initial search yielding 595 records (595 online databases, 9 from other sources), we removed 26 duplicates and screened the remaining 578 by title/abstract and full text. Eight studies met our inclusion criteria and were analyzed [8,17–19,32–35] (Fig. 1).

**Fig. 1.**
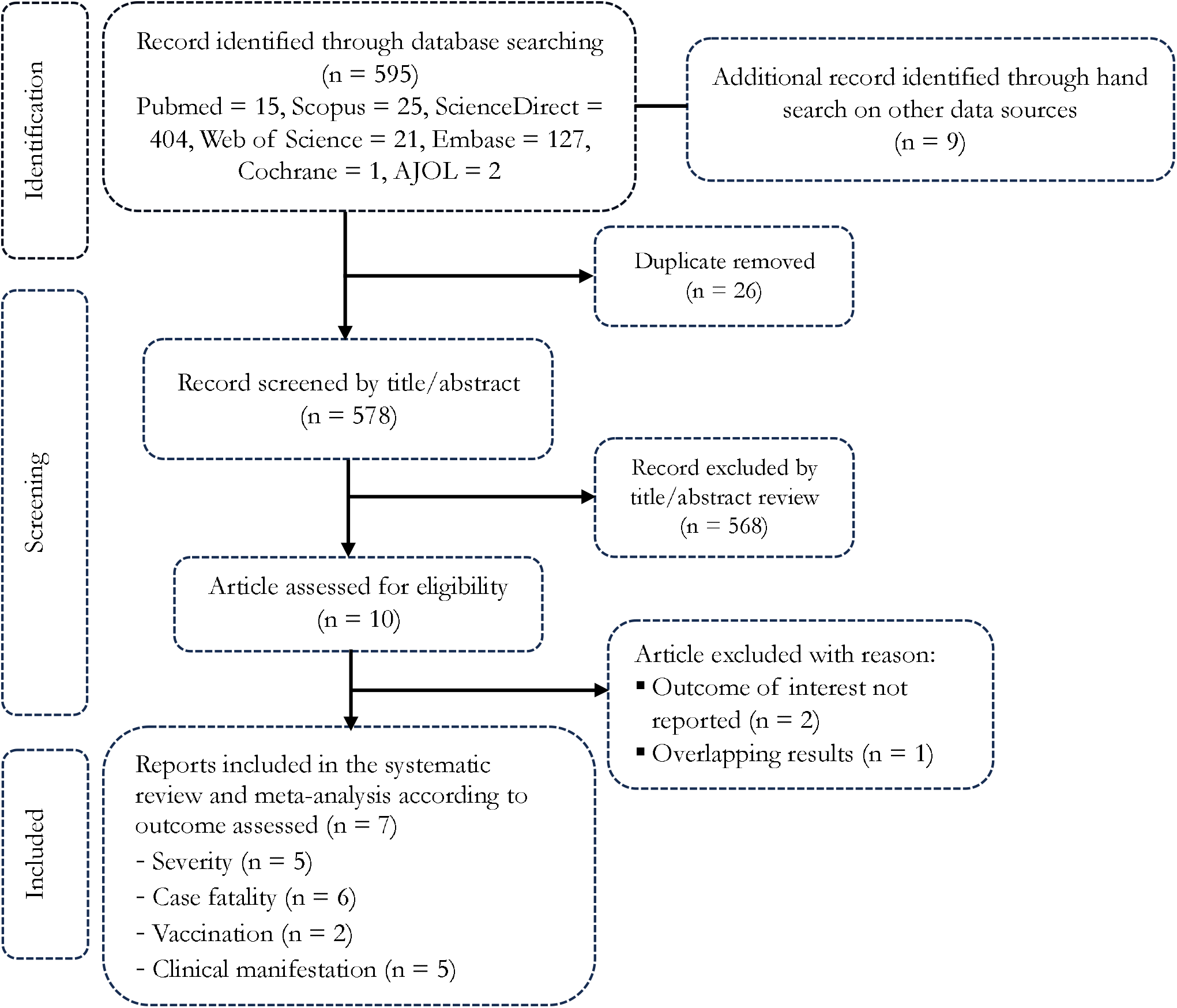
PRISMA diagram flow from study identification to inclusion in the meta-analysis

## Studies selection

### Characteristic of reports included

A comprehensive analysis included 7 studies. Those reports findings dated back from 1984 to 2023 and involved population from community and hospital settings. The studies were conducted nationally or in various regions from CAR including the Region 2, 4 and 6. All studies used surveillance and investigation methodology for data collection and a non-probabilistic sampling technique (Additional File 1, Supplementary Table 1)

### Disease severity among confirmed Mpox cases

The overall pooled Mpox severity rate among confirmed cases was 60.92% (95% confidence interval (CI): 47.54-72.83; *I*^2^ = 9.7% and *p* = 0.351) (Fig. 2).

**Fig. 2.**
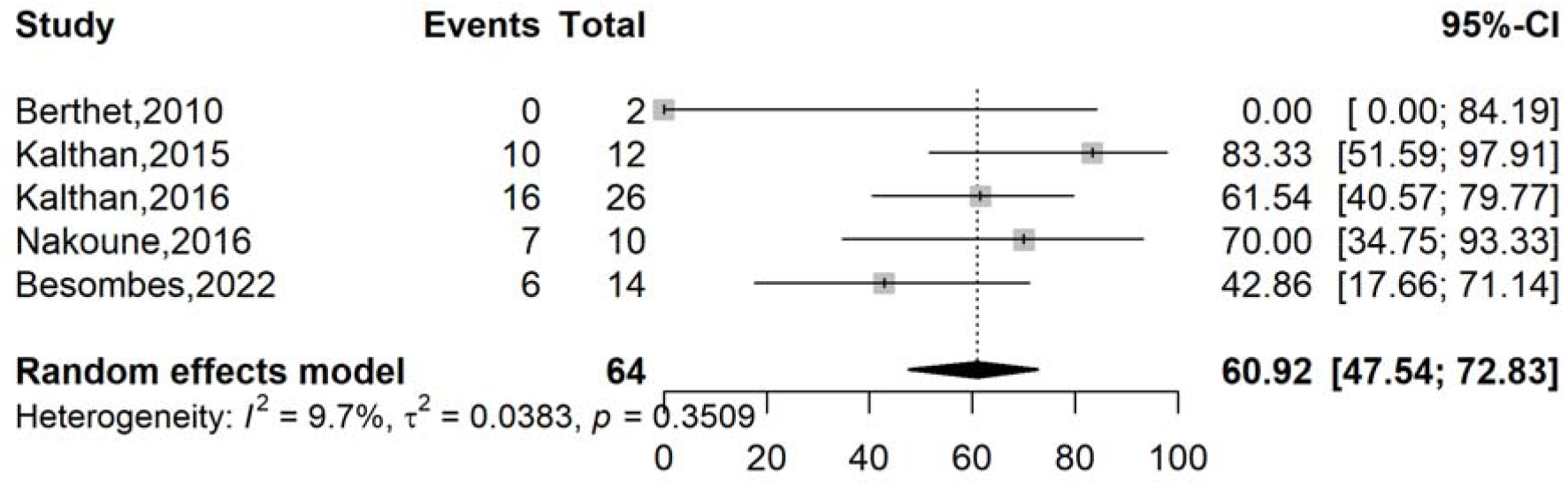
Pooled severity rate of Mpox cases in CAR

The subgroup analysis according to the study period revealed a non-significant decrease in Mpox severity rate from 66.00% (95% CI: 51.95-77.70; n = 4 reports) before 2022 [17–19,33] to 42.86 (95% CI: 17.66-71.14; n = 1 reports) [8]. The highest severity rate was observed in Region 6 (77.27%; 95% CI: 55.64-90.21; n = 2 reports) [17,33], however there was no significant difference between severity rate across regions (*p* = 0.120) (Table 1 and Additional Files 2, Supplementary Fig. 1-3).

**Table 1.** Subgroup meta-analysis of confirmed Mpox severity rate pooled estimates in CAR

| Subgroup | Confirmed cases | Event rate<sup>1</sup> (%) | 95% CI limits <sup>1</sup> |  | Number of studies | Heterogeneity statistic<sup>1</sup> |  |
| --- | --- | --- | --- | --- | --- | --- | --- |
|  |  |  | Lower | Upper |  | I<sup>2</sup>(%) | p-value |
| Period <sup>2</sup> |  |  |  |  |  |  |  |
| □ 2022 | 50 | 66.00 | 51.95 | 77.70 | 4 | 0 | 0.630 |
| 2022+ | 14 | 42.86 | 17.66 | 71.14 | 1 | - | - |
| Region |  |  |  |  |  |  |  |
| Region 2 | 14 | 42.86 | 17.66 | 71.14 | 1 | - | - |
| Region 4 | 26 | 61.54 | 40.57 | 79.77 | 1 | - | - |
| Region 6 | 22 | 77.27 | 55.64 | 90.21 | 2 | 0 | 0.463 |
| Not specified | 2 | 0.00 | 0.00 | 84.19 | 1 | - | - |
| **Participant** |  |  |  |  |  |  |  |
| General population | 54 | 59.26 | 45.81 | 71.45 | 4 | 26.6 | 0.252 |
| General population and HCW | 10 | 70 | 34.75 | 93.33 | 1 | - | - |

<sup>1</sup>Fixed effects model; CI: Confidence interval; <sup>2</sup>Period before and after the 2022 global outbreak; HCW: Healthcare worker

### Mortality among confirmed Mpox cases

The pooled CFR among confirmed Mpox cases was 11.54% (95% CI: 6.11-20.71; *I*^2^ = 0.0% and *p* = 0.893) (Fig. 3).

**Fig. 3.**
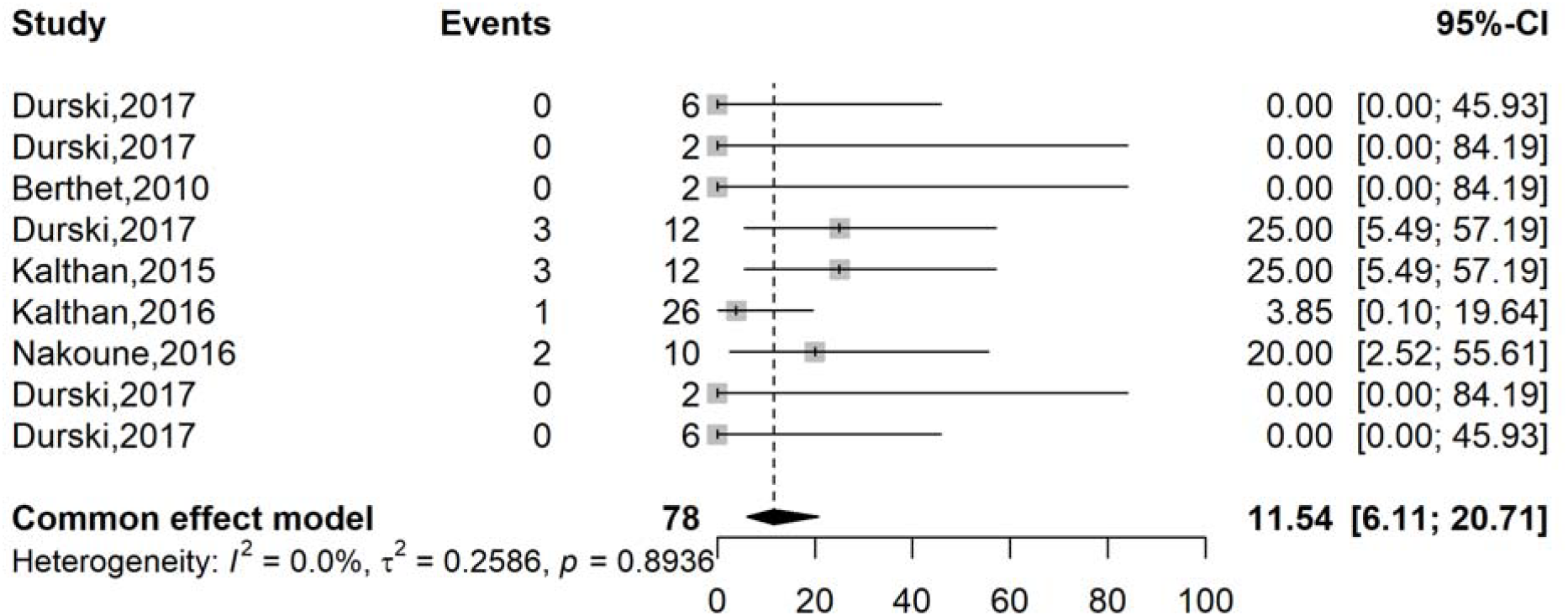
Pooled case fatality rate among confirmed Mpox cases in CAR

All included studies were conducted in before the global outbreak. The geographical trend highlighted higher mortality was recorded in Eastern Health Regions (11.11%; 95% CI: 5.08-22.60) [17,19,32,33] and the lowest in Western Health Regions with no reported death (0.00%; 95% CI: 0.00-100.00) [32] (Table 2 and Additional Files 3, Supplementary Fig. 1-3).

**Table 2.** Subgroup meta-analysis of confirmed Mpox case fatality rate pooled estimates in CAR

| Subgroup | Suspected cases | Event rate<sup>1</sup> (%) | 95% CI limits <sup>1</sup> |  | Number of studies | Heterogeneity statistic<sup>1</sup> |  |
| --- | --- | --- | --- | --- | --- | --- | --- |
|  |  |  | Lower | Upper |  | I<sup>2</sup>(%) | p-value |
| Region |  |  |  |  |  |  |  |
| Western | 8 | 0.00 | 0.00 | 100.00 | 2 | 0.0 | 1.000 |
| Eastern | 54 | 11.11 | 5.08 | 22.60 | 4 | 5 | 0.368 |
| Multicentric/National | 26 | 15.38 | 5.90 | 34.54 | 2 | 0.0 | 0.235 |
| Not specified | 4 | 0.00 | 0.00 | 100.00 | 2 | - | - |
| Participant |  |  |  |  |  |  |  |
| General population | 82 | 9.76 | 4.95 | 18.31 | 9 | 0.0 | 0.809 |
| General population and HCW | 10 | 20.00 | 2.52 | 55.61 | 1 | - | - |

<sup>1</sup>Fixed effects model; CI: Confidence Interval; Western = Health Regions 1 and 2; Eastern = Health Regions 4 and 6; HCW: Healthcare worker

### Mortality among suspected Mpox cases

The pooled CFR among suspected Mpox cases was 12.77% (95% CI: 7.39-21.15; *I*^2^ = 0.0% and *p* = 0.897) (Fig. 4).

**Fig. 4.**
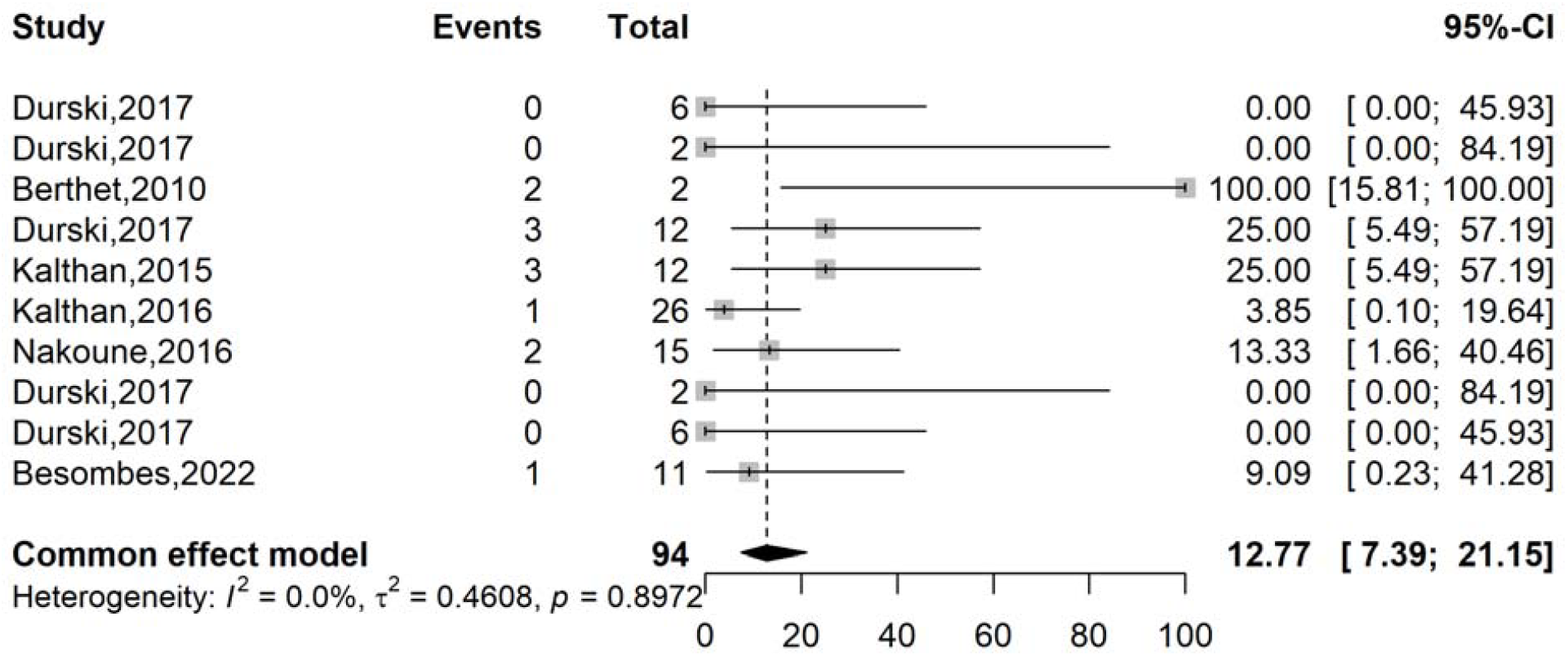
Pooled case fatality rate among suspected Mpox cases in CAR

The subgroup analysis shows a modest decline in case fatality rate (CFR) among suspected Mpox cases from the pre-2022 period (13.25%, 95% CI: 7.49-22.37) [17–19,32,33] to the post-2022 period (9.09%, 95% CI: 0.23-41.28) [35]. Geographic disparities existed, with Eastern Health Regions maintaining higher CFR (10.17%, 95% CI: 4.64-20.84) compared to Western Health Regions (5.26%, 95% CI: 0.74-29.39). The general population CFR (10.40%, 95% CI: 6.14-17.09) [8,18,19,32,33] aligns with overall trends, with no significant heterogeneity detected (*I*^2^ = 0.0-1.6%) across subgroups (Table 3 and Additional Files 4, Supplementary Fig. 1-4).

**Table 3.**
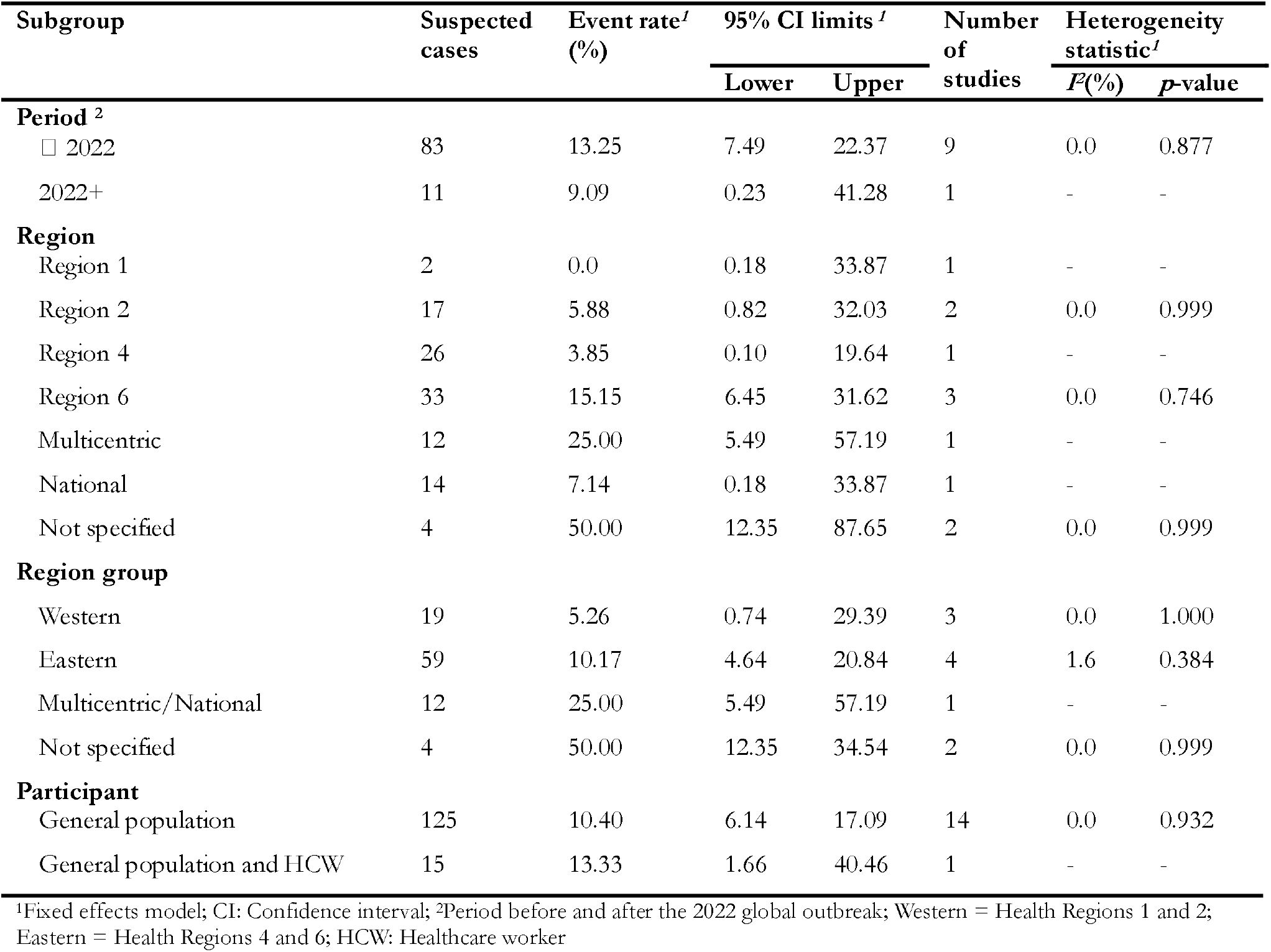
Subgroup meta-analysis of suspected Mpox case fatality rate pooled estimates in CAR

| Subgroup | Suspected cases | Event rate<sup>1</sup> (%) | 95% CI limits<sup>1</sup> |  | Number of studies | Heterogeneity statistic<sup>1</sup> |  |
| --- | --- | --- | --- | --- | --- | --- | --- |
| | | | Lower | Upper | | $I^2$(%) | p-value |
| **Period<sup>2</sup>** |  |  |  |  |  |  |  |
| □ 2022 | 83 | 13.25 | 7.49 | 22.37 | 9 | 0.0 | 0.877 |
| 2022+ | 11 | 9.09 | 0.23 | 41.28 | 1 | - | - |
| **Region** |  |  |  |  |  |  |  |
| Region 1 | 2 | 0.0 | 0.18 | 33.87 | 1 | - | - |
| Region 2 | 17 | 5.88 | 0.82 | 32.03 | 2 | 0.0 | 0.999 |
| Region 4 | 26 | 3.85 | 0.10 | 19.64 | 1 | - | - |
| Region 6 | 33 | 15.15 | 6.45 | 31.62 | 3 | 0.0 | 0.746 |
| Multicentric | 12 | 25.00 | 5.49 | 57.19 | 1 | - | - |
| National | 14 | 7.14 | 0.18 | 33.87 | 1 | - | - |
| Not specified | 4 | 50.00 | 12.35 | 87.65 | 2 | 0.0 | 0.999 |
| **Region group** |  |  |  |  |  |  |  |
| Western | 19 | 5.26 | 0.74 | 29.39 | 3 | 0.0 | 1.000 |
| Eastern | 59 | 10.17 | 4.64 | 20.84 | 4 | 1.6 | 0.384 |
| Multicentric/National | 12 | 25.00 | 5.49 | 57.19 | 1 | - | - |
| Not specified | 4 | 50.00 | 12.35 | 34.54 | 2 | 0.0 | 0.999 |
| **Participant** |  |  |  |  |  |  |  |
| General population | 125 | 10.40 | 6.14 | 17.09 | 14 | 0.0 | 0.932 |
| General population and HCW | 15 | 13.33 | 1.66 | 40.46 | 1 | - | - |

<sup>1</sup>Fixed effects model; CI: Confidence interval; <sup>2</sup>Period before and after the 2022 global outbreak; Western = Health Regions 1 and 2; Eastern = Health Regions 4 and 6; HCW: Healthcare worker

### Mpox vaccine uptake

The pooled vaccination uptake estimate was 20.00% (95% CI: 10.33-35.17) in CAR (Fig. 5)

**Fig. 5.**
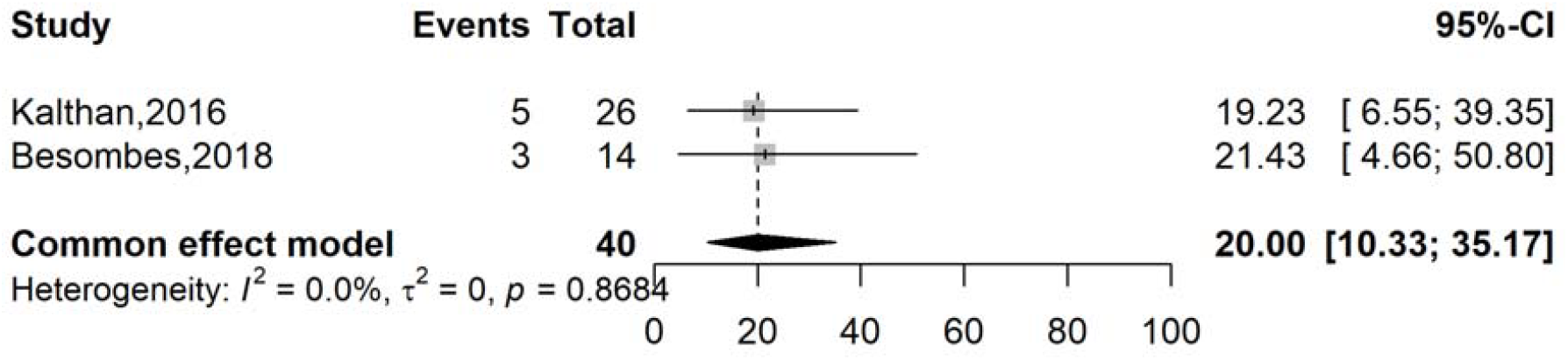
Pooled Mpox vaccination uptake rate in CAR

### Mpox clinical pattern

The most reported clinical manifestations associated with confirmed Mpox disease included fever, chills or sweat, light sensitivity, and rash with pooled frequencies varying from 91.05 to 85.53% of cases [8,17–19,33,35] (Table 4 and Fig. 6).

**Table 4.** Clinical patterns of confirmed Mpox cases in CAR

| Rank | Clinical manifestation | Confirmed cases<br>examined (n) | Frequency<br>(%) | 95% CI Limits |  | k | Heterogeneity<br>statistic |  | Model |
| --- | --- | --- | --- | --- | --- | --- | --- | --- | --- |
|  |  |  |  | Lower | Upper |  | *I*<sup>2</sup> (%) | *p*-value |  |
| 1 | Fever | 70 | 91.05 | 42.88 | 99.28 | 5 | 74.1 | 0.004 | Random |
| 2 | Chills or sweat | 10 | 90.00 | 55.50 | 99.75 | 1 | - | - | - |
| 3 | Light sensitivity | 9 | 88.89 | 51.75 | 99.72 | 1 | - | - | - |
| 4 | Rash | 76 | 85.53 | 75.72 | 91.80 | 6 | 15.8 | 0.312 | Fixed |
| 5 | Oral lesions | 20 | 59.16 | 8.56 | 95.73 | 2 | 86.5 | 0.007 | Random |
| 6 | Pruritus or itchy lesion | 46 | 58.57 | 31.53 | 81.27 | 3 | 59.1 | 0.087 | Random |
| 7 | Genital lesions | 19 | 57.43 | 8.76 | 94.99 | 2 | 85.4 | 0.009 | Random |
| 8 | Lymphadenopathy | 58 | 57.00 | 34.27 | 77.12 | 4 | 57.2 | 0.072 | Random |
| 9 | Fatigue or asthenia | 20 | 55.00 | 33.62 | 74.68 | 2 | 0.0 | 0.999 | Fixed |
| 10 | Palm lesions | 20 | 50.00 | 3.03 | 96.97 | 2 | 88.5 | 0.003 | Random |
| 11 | Sole lesions | 19 | 48.36 | 3.12 | 96.46 | 2 | 87.8 | 0.004 | Random |
| 12 | Lesions | 12 | 41.67 | 15.17 | 72.33 | 1 | - | - | - |
| 13 | Headache | 45 | 39.22 | 7.56 | 83.58 | 3 | 78.5 | 0.010 | Random |
| 14 | Sore throat or dysphagia | 35 | 37.87 | 11.67 | 73.76 | 2 | 83.5 | 0.014 | Random |
| 15 | Myalgia | 45 | 37.78 | 24.94 | 52.59 | 3 | 0.0 | 0.5770 | Fixed |
| 16 | Cough | 36 | 34.18 | 5.91 | 81.10 | 2 | 89.7 | 0.002 | Random |
| 17 | Conjunctivitis | 20 | 31.14 | 6.38 | 75.00 | 2 | 77.4 | 0.035 | Random |
| 18 | Hemorrhagic skin<br>lesions | 9 | 22.22 | 2.81 | 60.01 | 1 | - | - | - |
| 19 | Bedridden status | 10 | 20.00 | 2.52 | 55.61 | 1 | - | - | - |
| 20 | Vomiting or nausea | 10 | 20.00 | 2.52 | 55.61 | 1 | - | - | - |
| 21 | Diarrhea | 9 | 11.11 | 0.28 | 48.25 | 1 | - | - | - |
| 22 | Dyspnea | 10 | 10.00 | 0.25 | 44.50 | 1 | - | - | - |
| 23 | Facial oedema | 10 | 10.00 | 0.25 | 44.50 | 1 | - | - | - |
| 24 | Hypothermia | 10 | 10.00 | 0.25 | 44.50 | 1 | - | - | - |

k = Number of studies; CI: Confidence interval;

**Fig. 6.**
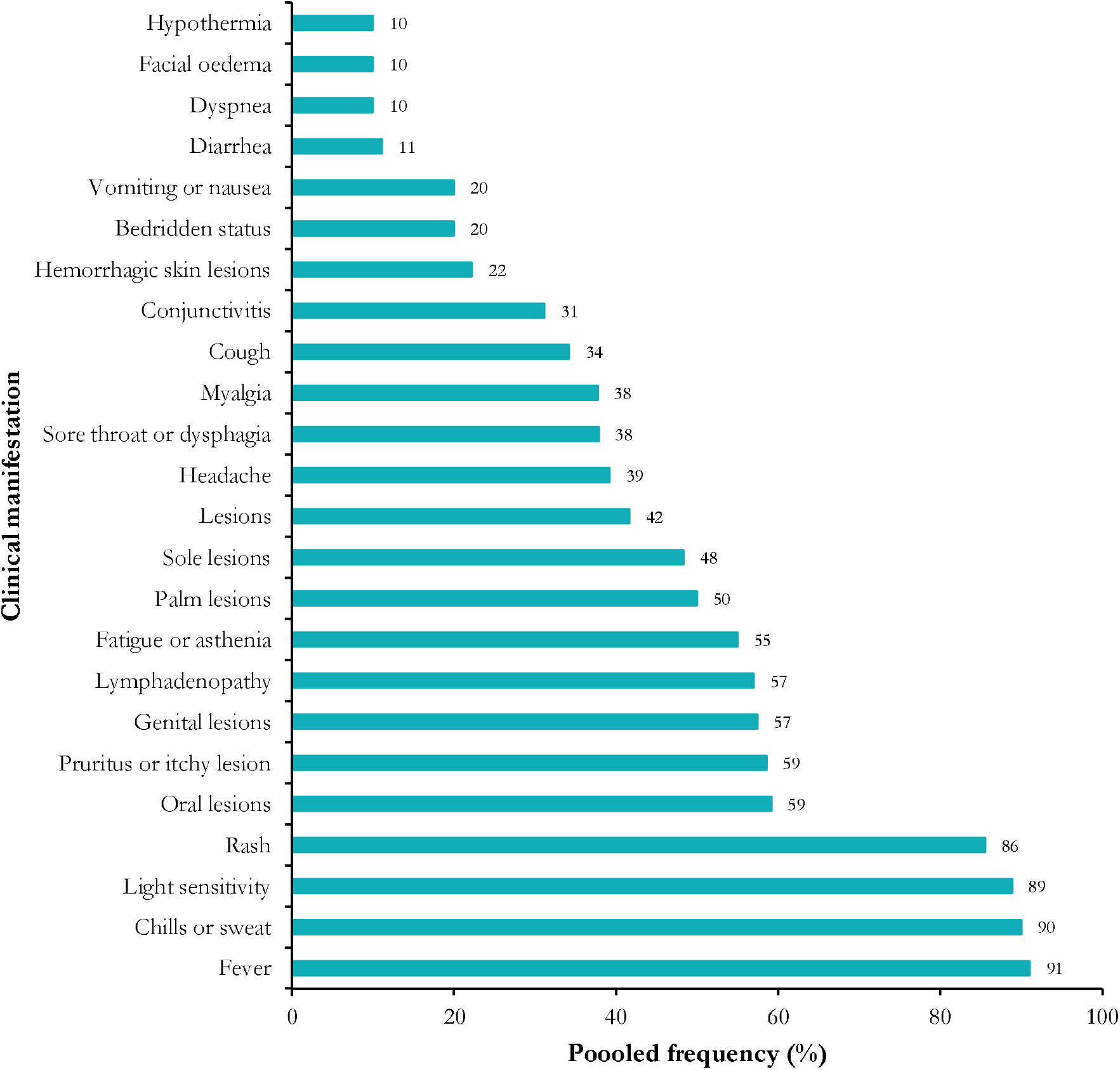
Clinical profile of confirmed Mpox cases in CAR

### Publication bias and sensitivity test analysis

A large inverted symmetrical distribution of data points was observed on the funnel plot for the four outcomes assessed, suggesting a low risk of publication bias for all the outcome interest. In addition, the Egger’s linear regression and Begg’s rank correlation tests confirmed the absence of statistically significant publication bias for the CFR among suspected cases (Additional Files 2, Supplementary Fig. 4; Additional Files 3, Supplementary Fig. 4; Additional Files 4, Supplementary Fig. 5; and Additional File 5 Supplementary Fig. 1).

Sensitivity analysis, assessing the impact of individual studies and outliers on the overall results, demonstrated that no study exhibited a significant impact on the overall pooled estimate (Additional Files 2, Supplementary Fig. 5; Additional Files 3, Supplementary Fig. 5; and Additional Files 4, Supplementary Fig. 5).

## Discussion

This in-depth systematic analysis and meta-analysis involved a comprehensive synthesis of 15 studies conducted in CAR between 1984 and 2023. These results provide critical insights into the epidemiological, clinical, and public health patterns of the Mpox virus in CAR.

### Case fatality and clinical severity

In this study, an observed CFR of 11.54% was reported among confirmed cases. This is markedly higher than figures reported in recent global outbreaks, where most studies reported CFR to remain consistently below 1%[36–38]. This high CFR is similar to earlier studies from the Democratic Republic of Congo (DRC), where surveillance between 2001 and 2013 revealed CFRs ranging from 6% to 10.6%, particularly in children and unvaccinated individuals [39,40]. Furthermore, similar mortality rates were documented in the CAR during earlier outbreaks in the 1980s [41]. The high CFR in CAR can be attributed to factors such as the endemic circulation of the more virulent clade I Mpox virus among CAR cases, limited healthcare access, poor health-seeking behavior, and frequent delays in case detection and isolation, especially in rural communities. This highlights the need for collaborative and resilient healthcare systems to reduce this mortality.

Notably, our analysis showed a higher CFRs among confirmed in the Eastern Health Regions (11.11%; 95% CI: 5.08–22.60) compared to the Western Regions (0.0%; 95% CI: 0.00–100.00), with elevated mortality particularly concentrated in localities reporting a higher burden of suspected cases. These areas overlap with regions most affected by the prolonged conflict following the 2013 arm conflict, such as Haute-Kotto, Vakaga, Mbomou, and Ouham [42]. These zones have experienced repeated armed violence, mass displacement, and targeted attacks on health facilities, severely compromising access to healthcare services. Médecins Sans Frontières and other humanitarian agencies have reported recurrent looting, destruction of infrastructure, and withdrawal of essential health services in conflict-affected health districts such as Batangafo, Bambari, and Bangassou [43]. The ongoing insecurity continues to restrict healthcare workers’ mobility and disrupt medical supply chains, further impeding timely diagnosis, isolation, and treatment of Mpox cases.

These conflict-driven vulnerabilities likely contribute significantly to the elevated mortality and underscore the urgent need for collaborative and resilient healthcare systems that can operate effectively amid political instability and violence to reduce Mpox-related deaths in CAR.

The Central African Republic’s extensive border with the Democratic Republic of Congo (DRC) - the regional epicenter of Mpox - represents a critical transmission risk, particularly in Health Region 6 where the highest disease severity rate was documented. This border region’s dense forest ecosystems and local populations’ dependence on hunting, gathering, and agricultural activities create frequent human-animal interactions that facilitate zoonotic spillover [44]. The combination of cross-border movement and traditional livelihoods in these forested zones likely explains the elevated Mpox transmission observed in these communities and subsequent likelihood of high severity in this setting.

About 61% of cases with clinical severity in CAR reported symptoms such as fever (91%), rash (86%), photophobia (89%), and fatigue (55%). These presentations are consistent with earlier descriptions of Clade I infections, which are known to be responsible for systemic illness with complications such as secondary bacterial infections, dehydration, and acute respiratory distress[40,45]. The WHO has previously characterized Clade I as causing more severe and widespread disease, with a higher propensity for complications and mortality compared to Clade IIa or IIb [46].

On the other hand, the 2022–2023 Mpox outbreaks, predominantly caused by Clade IIb, presented with milder disease profiles. An extensive multinational cohort study across 16 countries by Thornhill *et al.* found that most patients exhibited localized anogenital lesions with minimal systemic symptoms, and hospitalization was required in less than 10% of cases [37]. Similarly, Tarín-Vicente et al [36] observed that although pain was a common symptom, especially with perianal lesions, life-threatening complications were rare, and the CFR was negligible. This phenotypic difference reflects both virological differences and contextual health infrastructure disparities across many countries, especially in Africa, where timely access to quality and affordable healthcare remains limited in most countries.

Another major challenge in CAR is underreporting, which underestimates the actual burden of Mpox reported over the years. Rural communities often lack access to quality healthcare, including diagnostic testing and trained personnel. Studies have highlighted that several cases may go undetected for every confirmed Mpox case in endemic areas due to inadequate surveillance [47]. Moreover, symptoms of Mpox can overlap with other endemic conditions like varicella, measles, or bacterial skin infections, leading to misclassification and diagnostic delays [48]. The low testing and reporting rates hinder effective outbreak response, delay contact tracing, and impair understanding of virus evolution.

On the other hand, during recent outbreaks, a number of African nations have shown successful testing and reporting techniques. For example, Nigeria expanded molecular diagnostics through the Nigeria Centre for Disease Control during the 2022–2023 mpox outbreak, allowing for weekly situation reporting and prompt case confirmation [49]. Similarly, real-time reporting and trend monitoring of monkeypox have been enhanced in the Democratic Republic of the Congo through the implementation of surveillance technologies such as DHIS2 and systematic case notification [50].

### Vaccination access and global inequity

The review showed a low vaccination uptake in CAR (20%). These results corroborates finding from the neighboring DRC [39]. This could be due to supply limitations, healthcare infrastructural challenges, and vaccine hesitancy among the population. Historically, smallpox vaccination provided cross-protection against Mpox, with immunity estimates around 85% [51]. However, the cessation of global smallpox immunization in 1980 resulted in a growing cohort of susceptible individuals in endemic regions [52]. Countries like CAR, with weak immunization infrastructure, have since lacked routine smallpox-derived protection and access to new Mpox-specific vaccines.

In contrast, during the 2022 outbreak, developed countries rapidly deployed targeted vaccination campaigns. For example, the U.S. administered over 1.2 million doses of JYNNEOS by early 2023 [53], prioritizing healthcare workers and most vulnerable groups like men who have sex with men. Similarly, the UK initiated a pre-exposure prophylaxis campaign using a ring vaccination strategy [54]. These discrepancies showed broader global health inequities in global vaccine distribution efforts and research funding towards endemic countries like CAR, which the Mpox morbidity and mortality have impacted for decades.

### Strength and limitations

This systematic review and meta-analysis provide valuable insights into Mpox severity, mortality and the clinical profile of confirmed cases to tailor a context specific Mpox case definition in the CAR. The findings highlight the continued public health importance of Mpox in the region and underscore the need for ongoing surveillance, research on vaccination uptake, and interventions to reduce the burden of this disease. However, this review has several limitations. First, some death cases might have been unreported due to study design, potentially underestimating the true CFR. Confounding by strain differences remains a concern, as the lack of comprehensive genomic data in many studies obscures the clade-specific mortality rates. Surveillance bias, particularly the potential for improved detection of milder cases in more recent studies, may also influence the CFR estimates. Additionally, the small sample sizes in some subgroup analyses result in wide confidence intervals, limiting the precision of the findings. Study in other language than English and French were not searched.

## Conclusions

Mpox remains a major public health challenge in the Central African Republic, with mortality and disease severity far exceeding those reported in recent non-endemic outbreaks. This disparity is primarily due to the circulation of the more virulent Clade I virus and also reflects limited healthcare access, low vaccination coverage, and underinvestment in surveillance infrastructure. Future research in the country should explore and confirm these gaps in the management of Mpox cases. Consequently, the international response to Mpox must address these structural inequities through enhanced resource allocation, targeted vaccine delivery, community education, healthcare worker training, and research partnerships with endemic countries.

## Supporting information

Addition Files 4

Addition Files 5

Addition Files 1

## Data Availability

All data produced in the present work are contained in the manuscript and its supplementary materials

## Abbreviations

CAR: Central African Republic
CFR: Case Fatality Rate
CI: Confidence Interval
DRC: Democratic Republic of Congo
HCW: Healthcare worker
MeSH: Medical Subject Headings
PRISMA: Preferred Reporting Items for Systematic Reviews and Meta-Analysis
WHO: World Health Organization

## Declarations

### Author contributions

F.Z.L.C. conceived the original idea of the study; F.Z.L.C. and C.A. conducted the literature search; F.Z.L.C., D.R.T. and L.B.K.B. selected the studies, extracted the relevant information, and synthesized the data. F.Z.L.C. performed the analyses and wrote the first draft of the manuscript. All authors critically reviewed and revised successive drafts of the manuscript. All authors read and approved of the final manuscript.

### Ethical approval statement

Not applicable

### Consent for publication

Not applicable.

### Availability of data and materials

The sources of data supporting this systematic review are available in the reference. All data generated or analyzed during this study are included in this published article and supplemental material.

