## Additional Files 3 for "Mpox clinical and epidemiological patterns in the Central African Republic: a systematic review and meta-analysis"

**Region**

**Case fatality rate (%)**

**Event rate (%)**

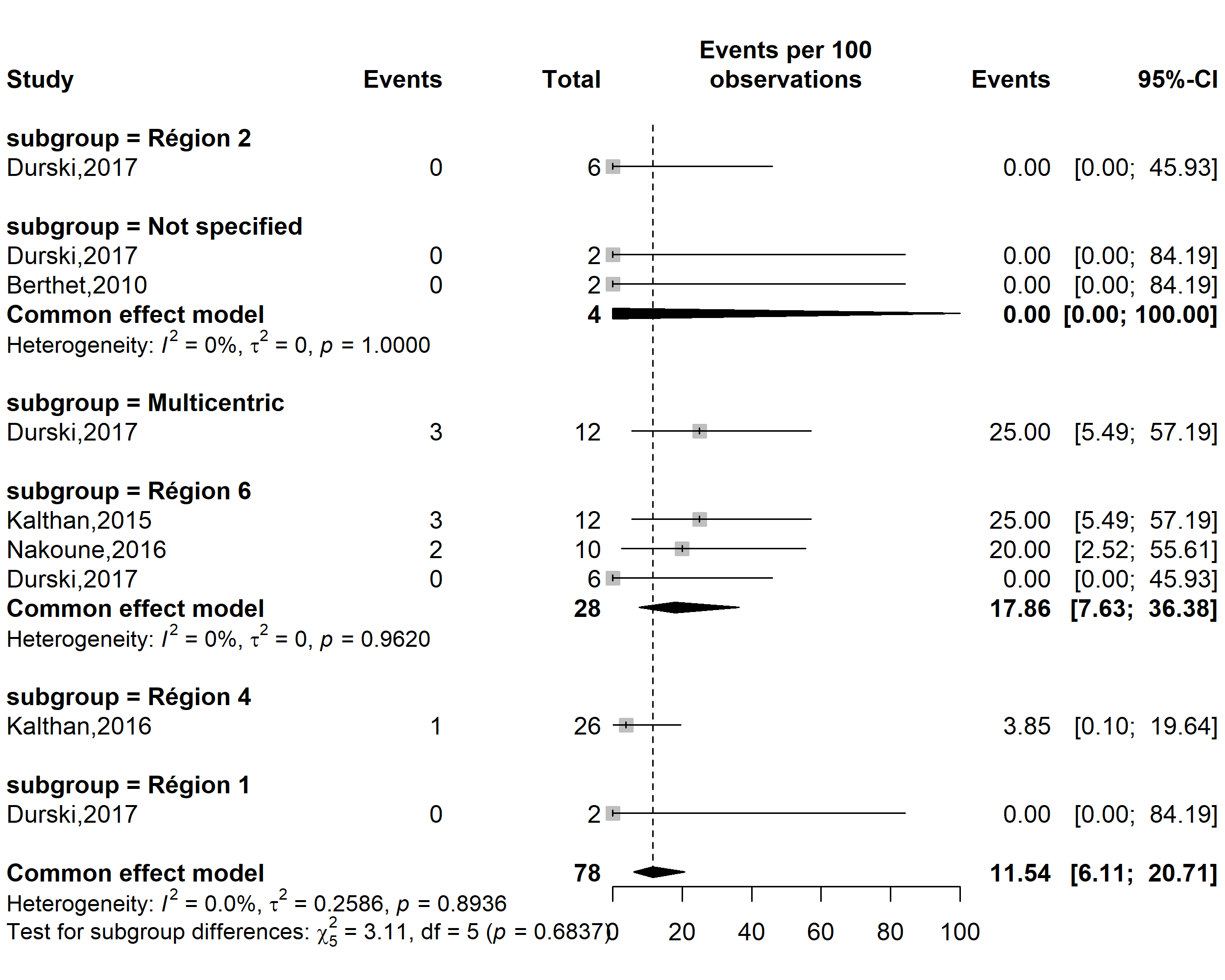

Supplementary Fig. 1 Pooled case fatality rate among confirmed Mpox cases by CAR’s regions

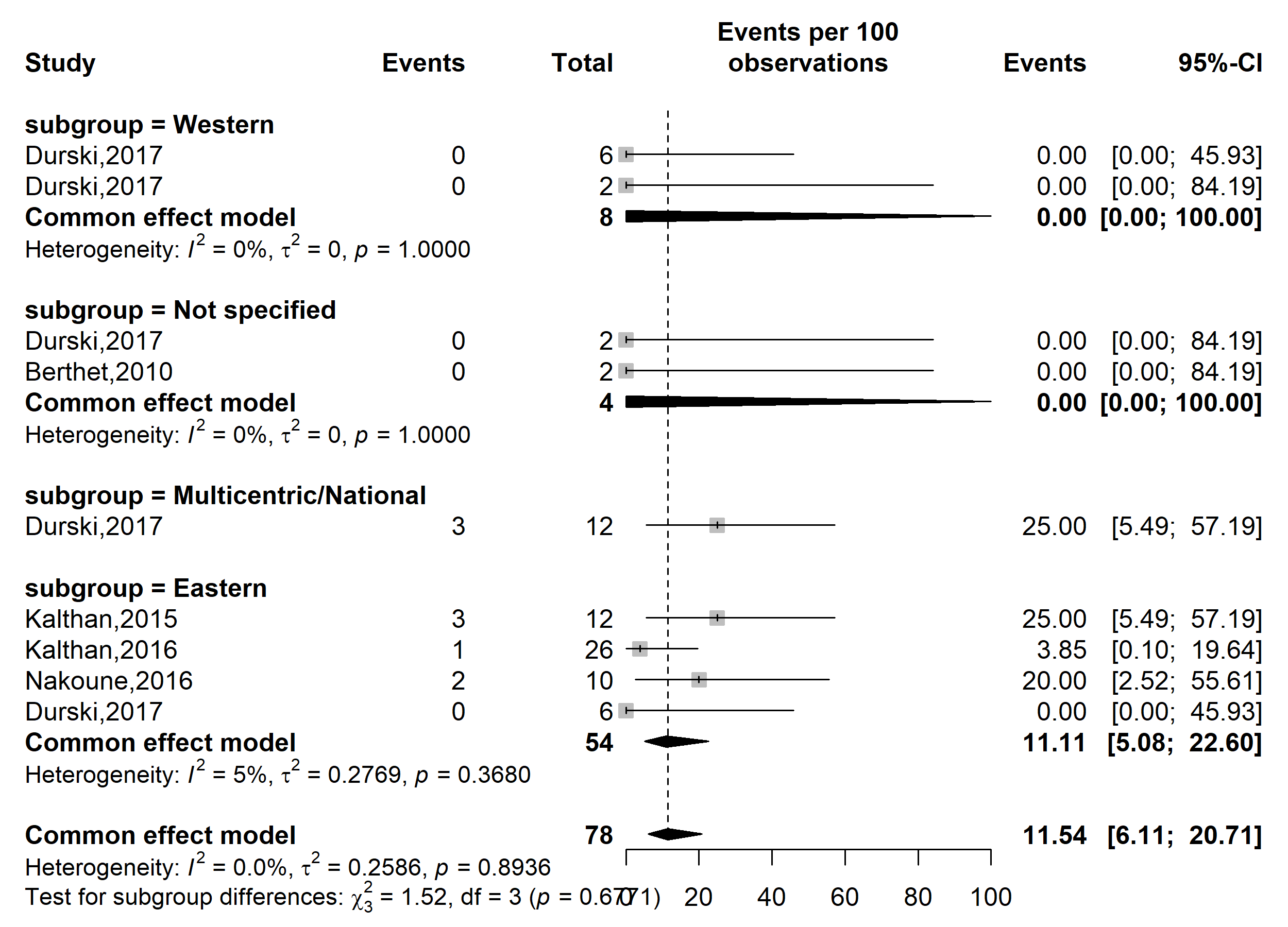

**Case fatality rate (%)**

**Event rate (%)**

Supplementary Fig. 2 Pooled case fatality rate by region location among confirmed Mpox cases in CAR *(HCW: Healthcare worker)*

**Participant**

**Case fatality rate (%)**

**Event rate (%)**

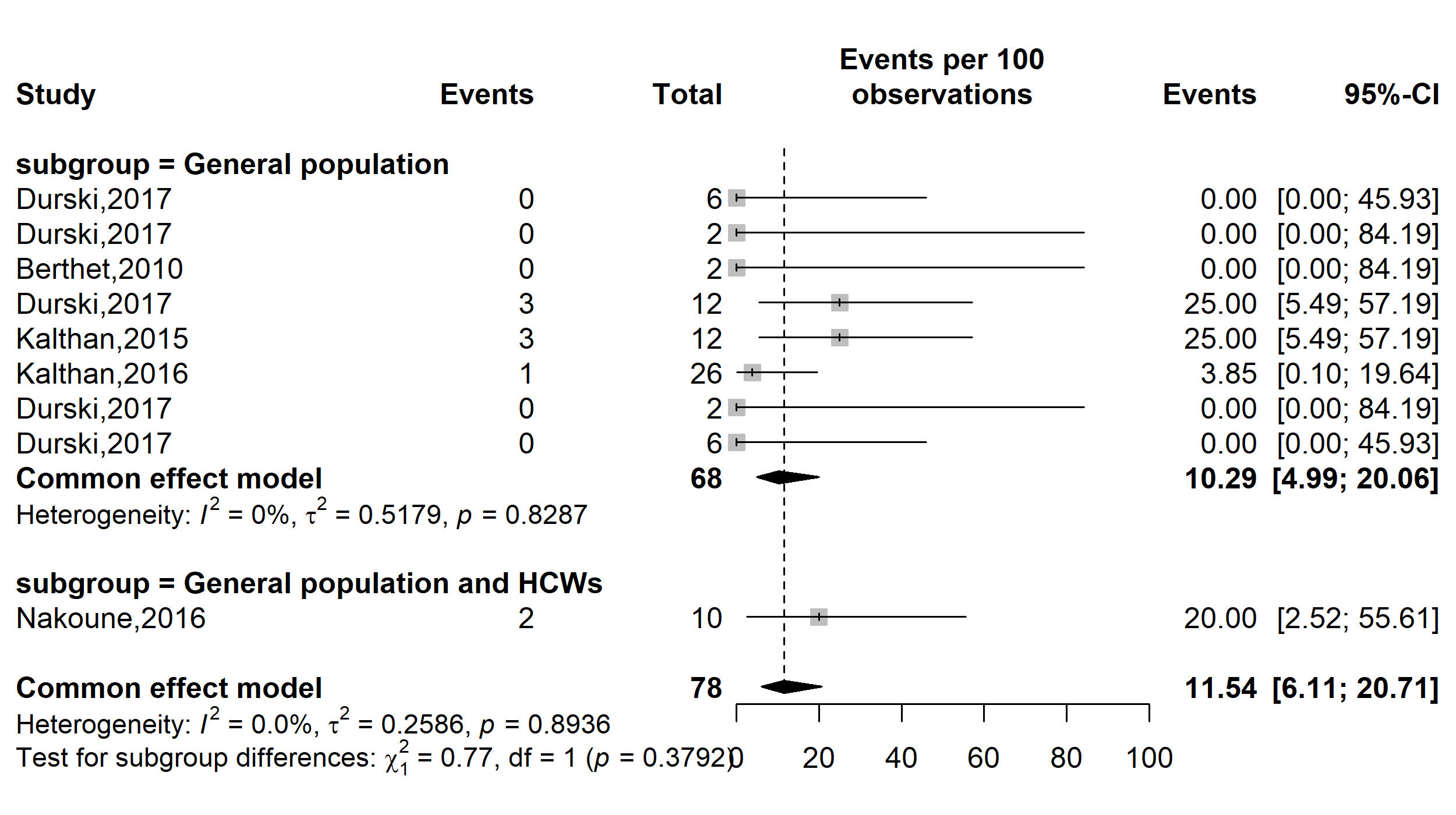

Supplementary Fig. 3 Pooled case fatality rate by type of participants involved among confirmed Mpox cases in CAR *(HCWs: Healthcare workers)*

**Publication bias**

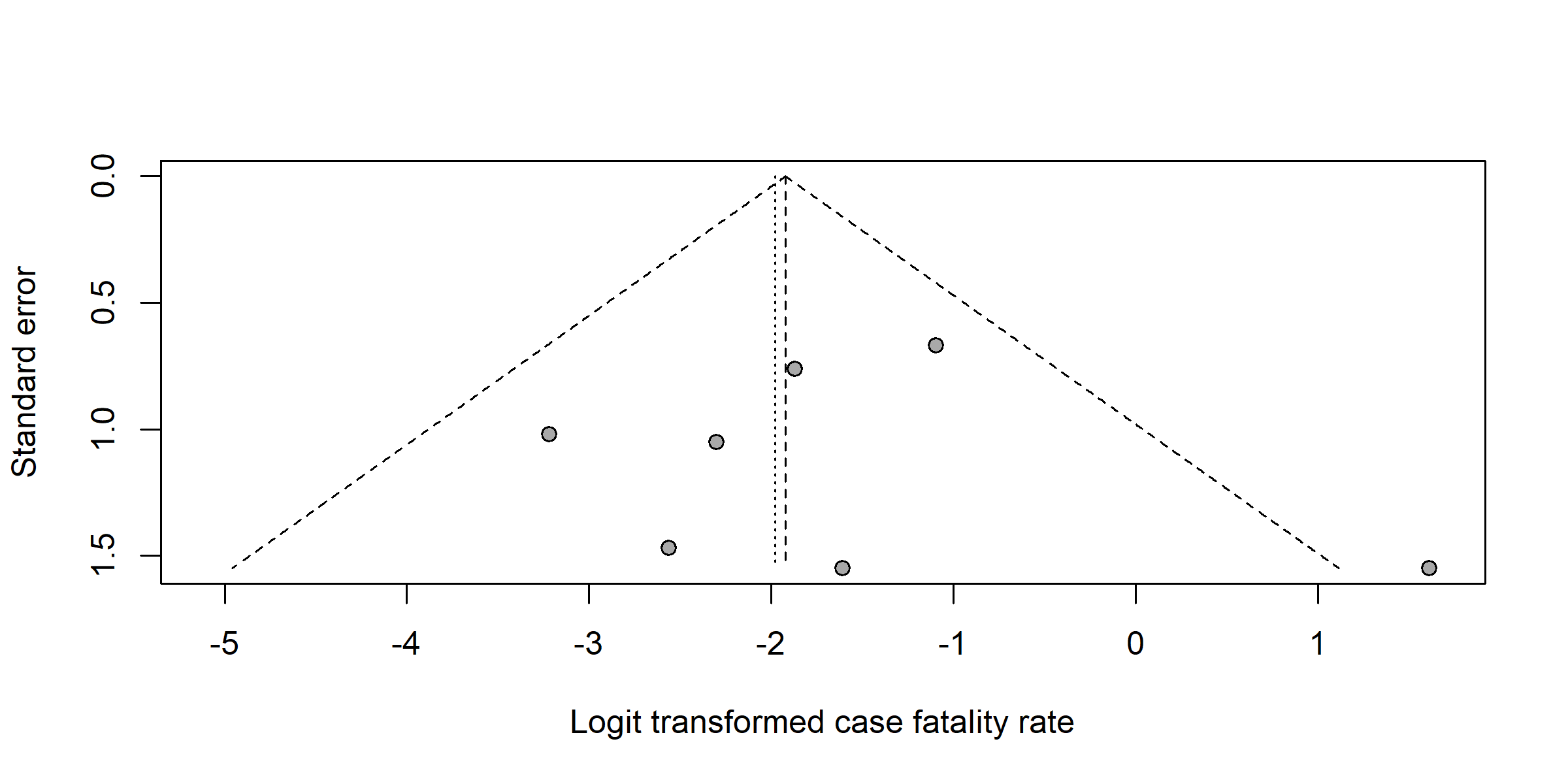

Supplementary Fig. 4 Funnel plot with pseudo 95% confidence limits and tests assessing the publication bias of studies included

**Sensitivity analysis**

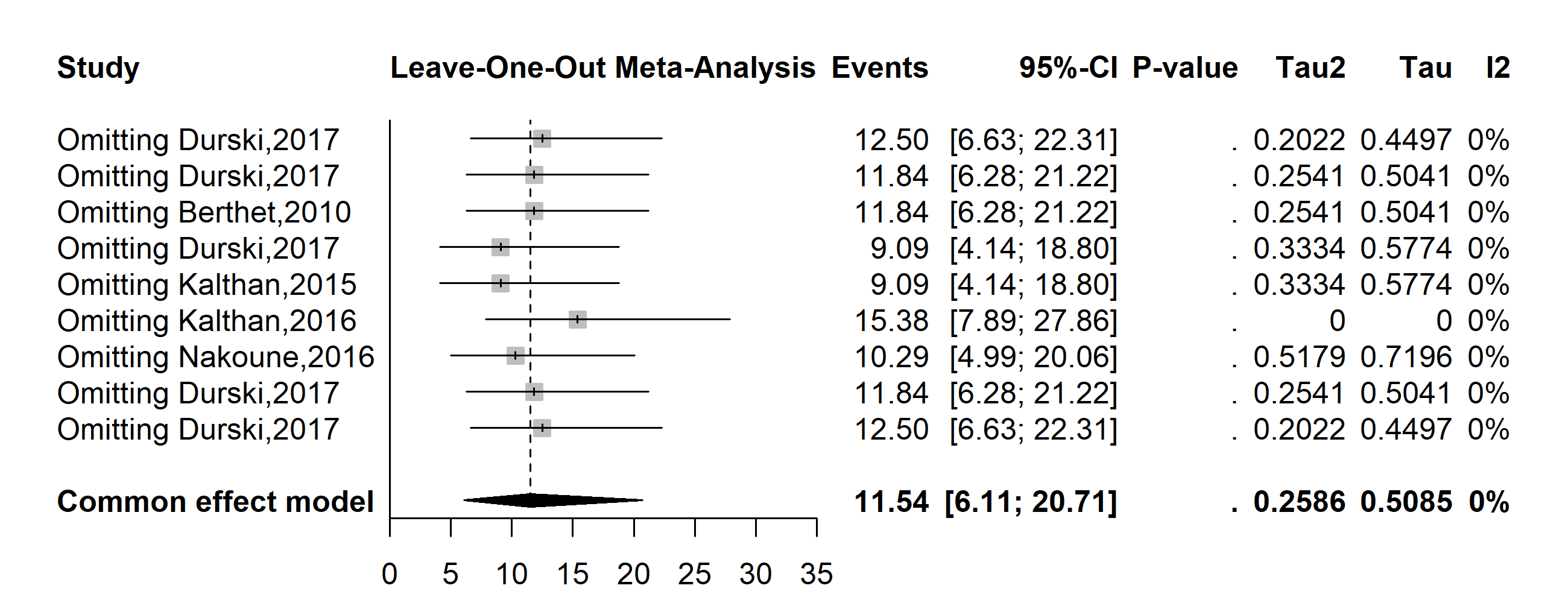

Supplementary Fig. 5 Sensitivity analysis of the confirmed Mpox case fatality rate pooled estimate in CAR
