## Supplementary material for "Mpox clinical and epidemiological patterns in the Central African Republic: a systematic review and meta-analysis": Addition Files 4

**Period**

**Event rate (%)**

**Case fatality rate (%)**

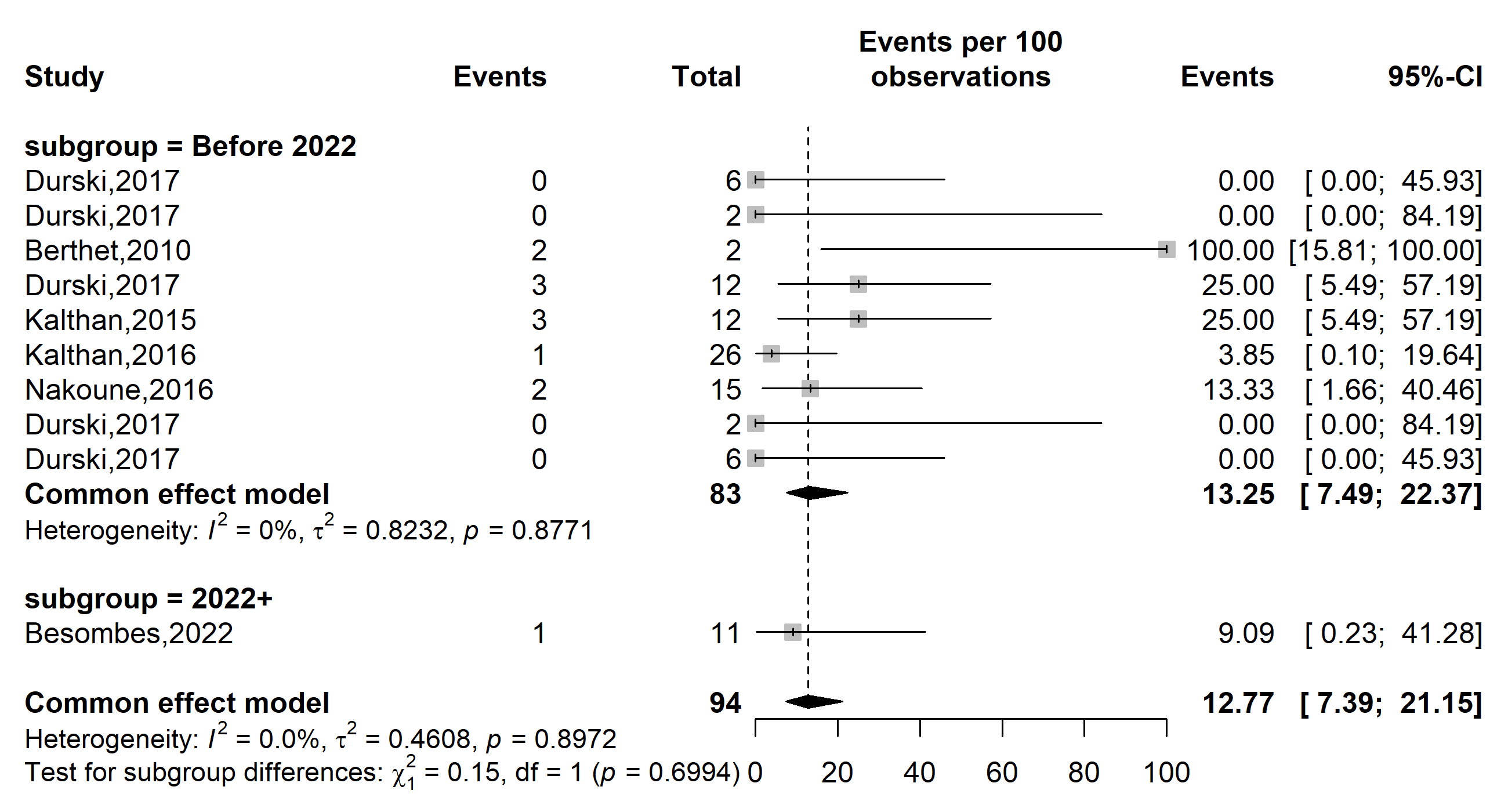

Supplementary Fig. 1 Pooled case fatality rate by study period among suspected Mpox cases in Central African Republic (CAR) (*the period represents before and since the 2022 pandemic*)

**Region**

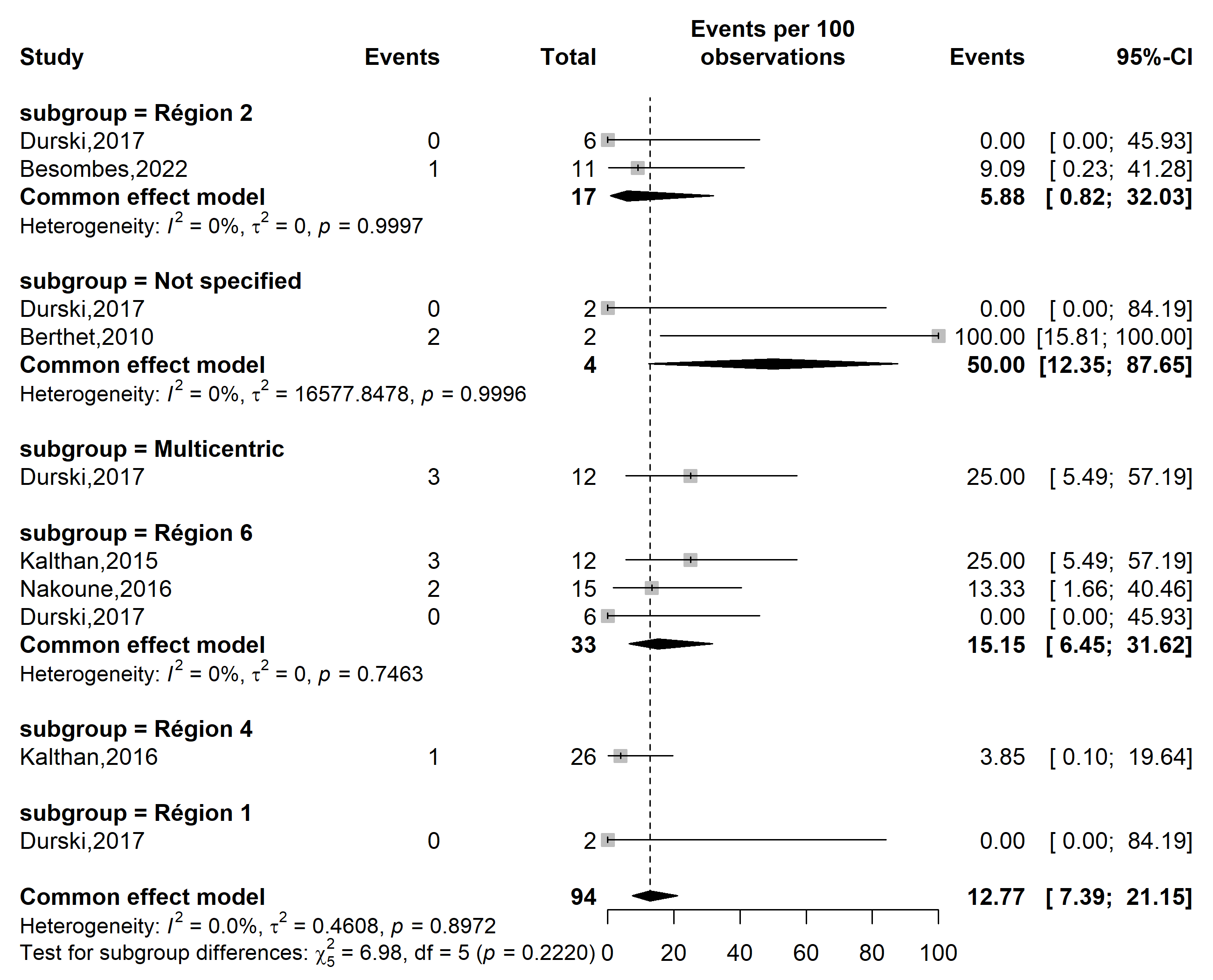

**Case fatality rate (%)**

**Event rate (%)**

Supplementary Fig. 2 Pooled case fatality rate among suspected Mpox cases by CAR’s regions

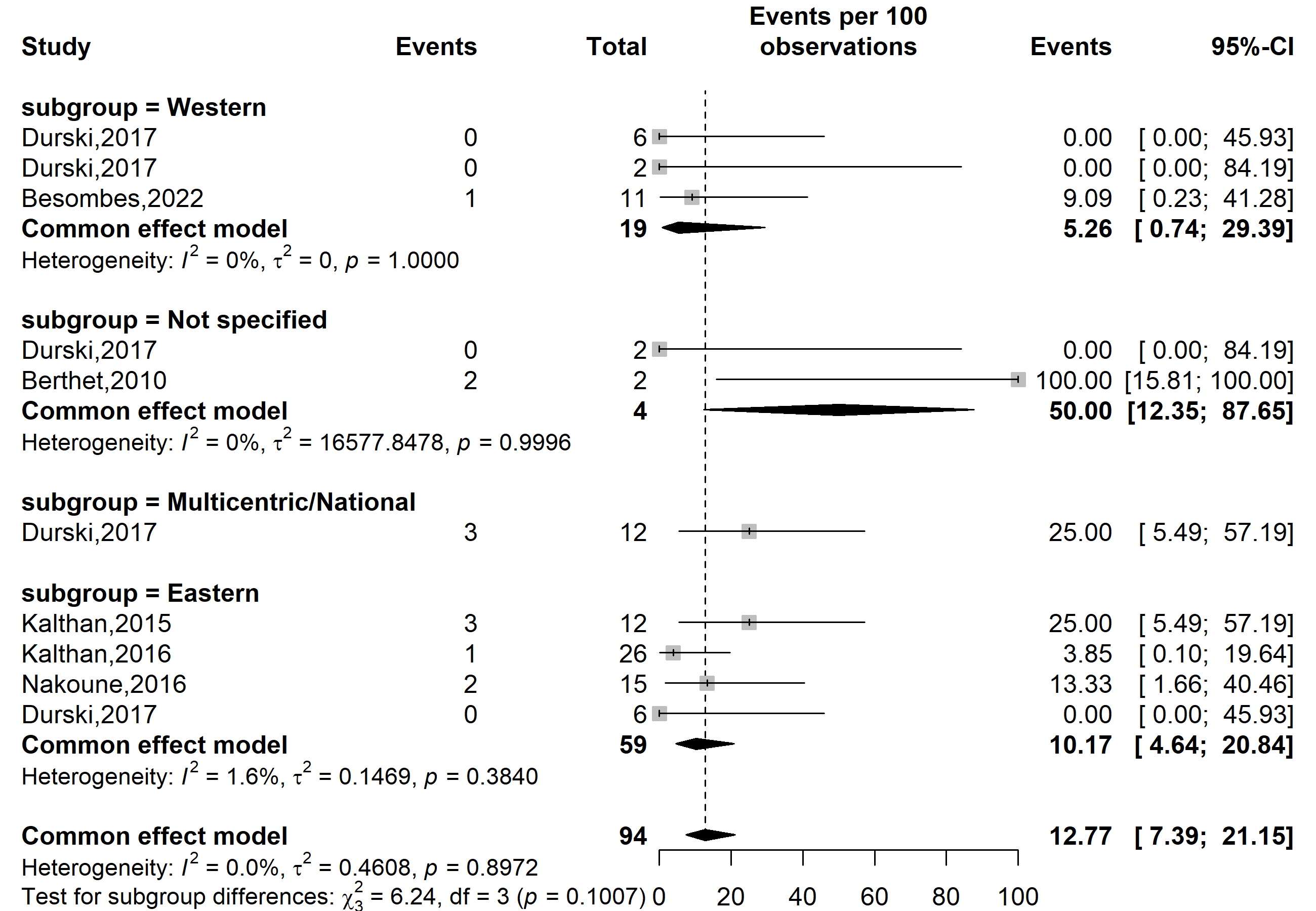

**Case fatality rate (%)**

**Event rate (%)**

Supplementary Fig. 3 Pooled case fatality rate by region location among suspected Mpox cases in CAR *(HCW: Healthcare worker)*

**Participant**

**Event rate (%)**

**Case fatality rate (%)**

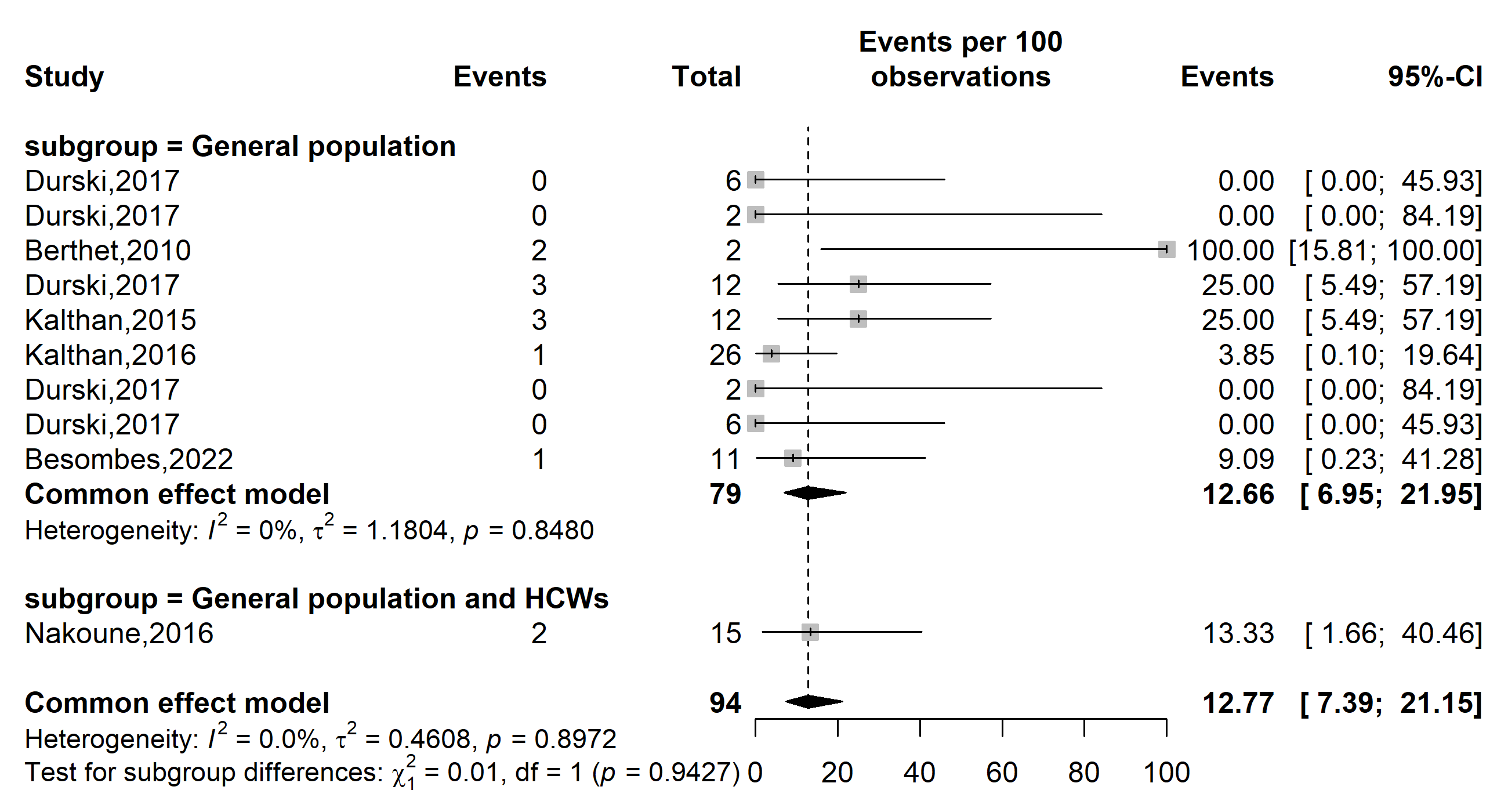

**Supplementary Fig. 4** Pooled case fatality rate by type of participants involved among suspected Mpox cases in CAR *(HCW: Healthcare worker)*

**Publication bias**

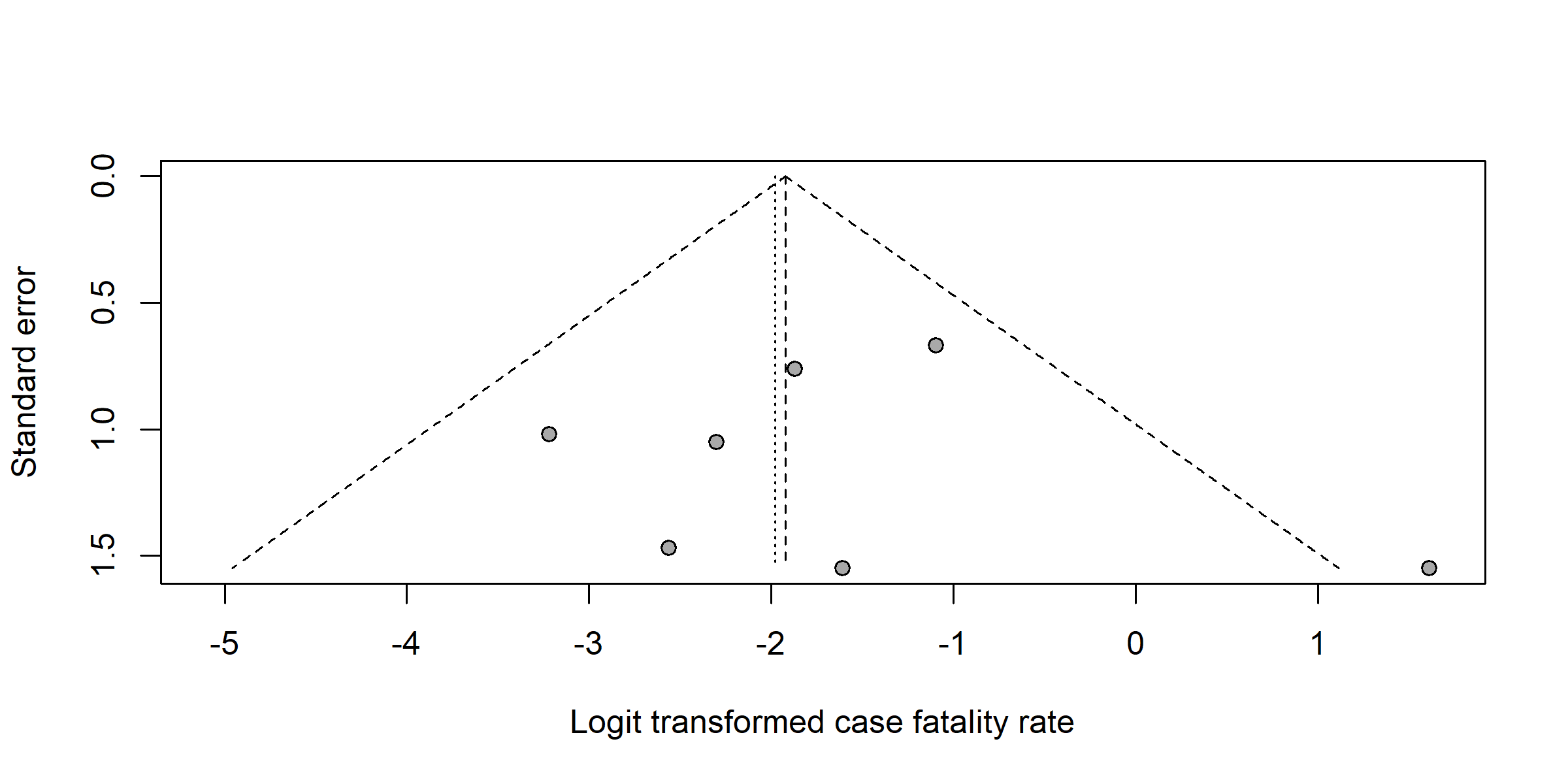

Egger’s test (*p*-value = 0.743)

Begg’s test (*p*-value = 0.711)

Supplementary Fig. 5 Funnel plot with pseudo 95% confidence limits and tests assessing the publication bias of studies included

**Sensitivity analysis**

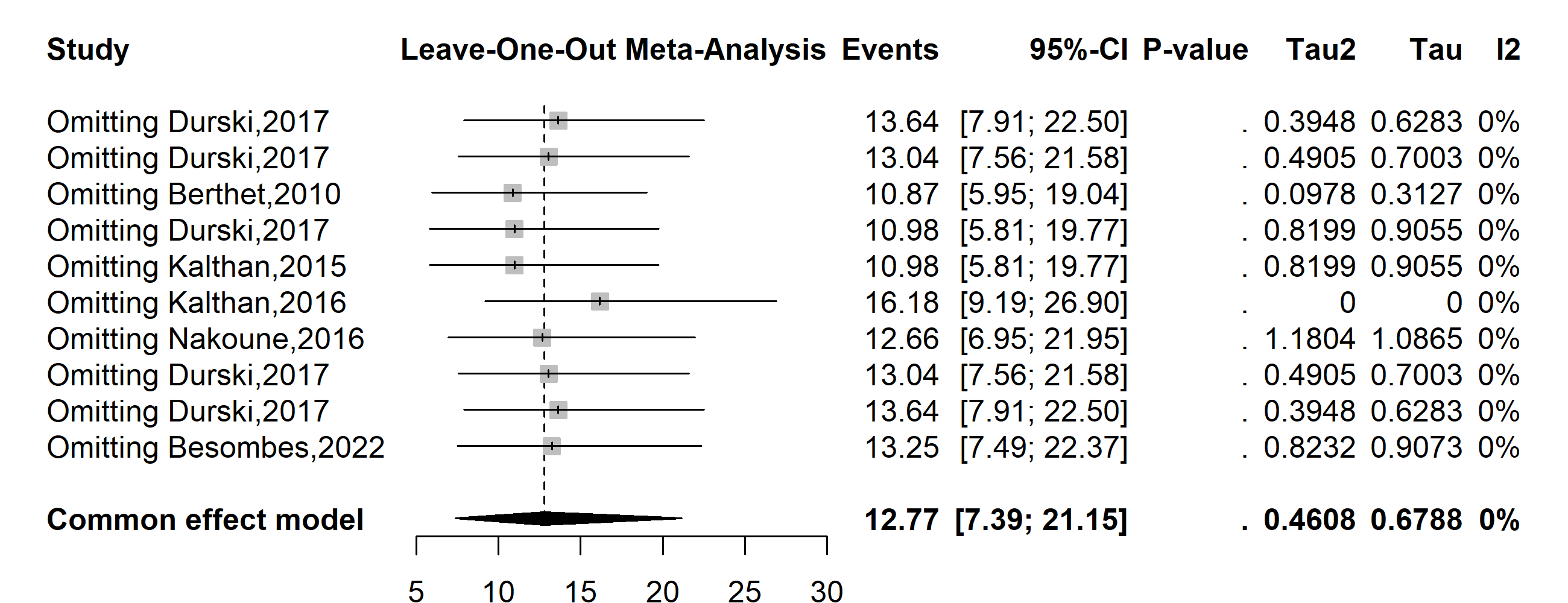

Supplementary Fig. 6 Sensitivity analysis of the suspected Mpox case fatality rate pooled estimate in CAR
