## Supplementary material for "Mpox clinical and epidemiological patterns in the Central African Republic: a systematic review and meta-analysis": Addition Files 5

**Publication bias**

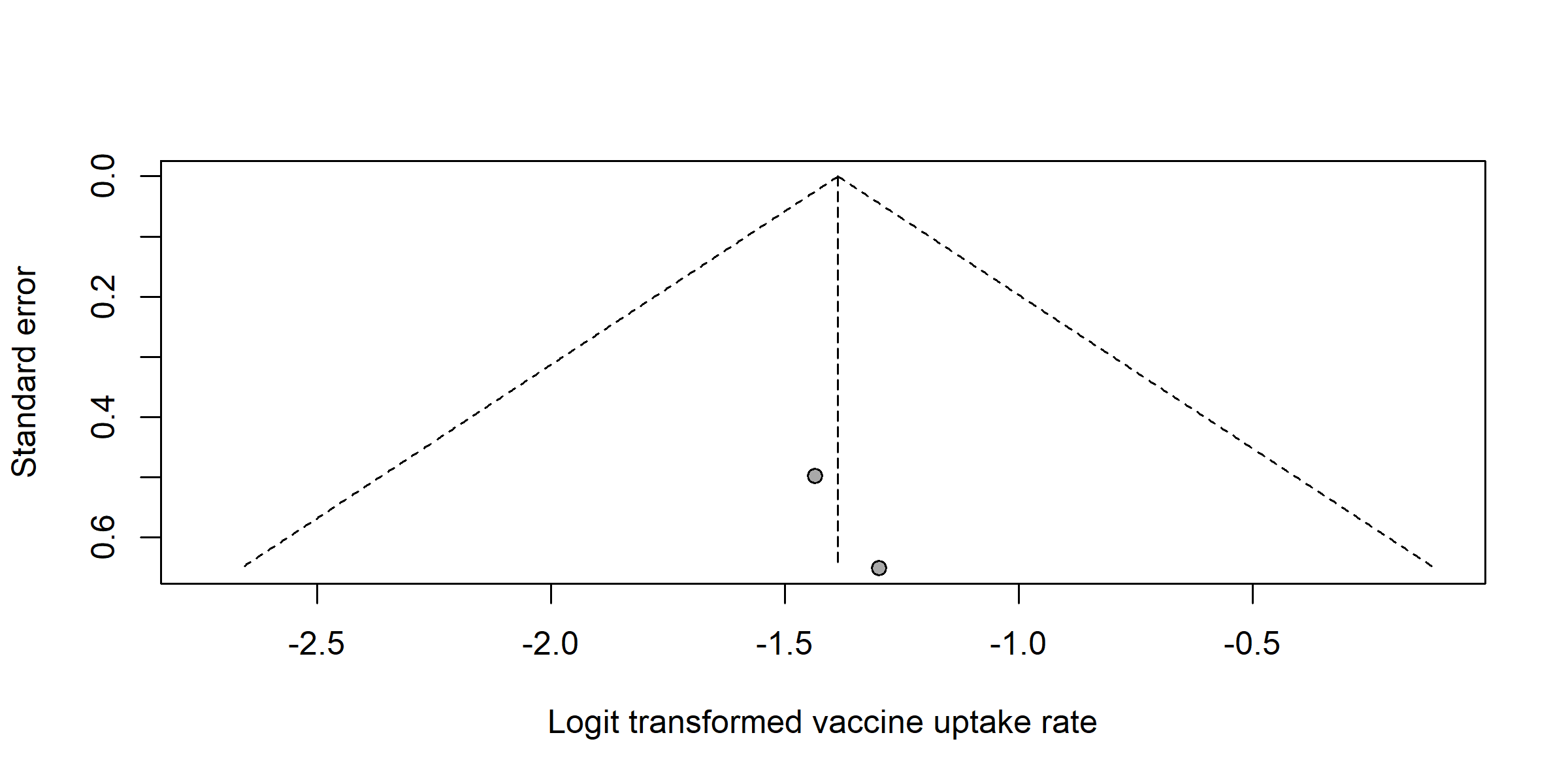

Supplementary Fig. 1 Funnel plot with pseudo 95% confidence limits and tests assessing the publication bias of studies included to pooled vaccination uptake.
