## Supplementary material for "Mpox clinical and epidemiological patterns in the Central African Republic: a systematic review and meta-analysis": Addition Files 1

Supplementary Table 1 Characteristic of studies assessing Mpox severity and mortality in DRC, 1970-2024

| Author | Study  Year*^1^* | Region | Setting | Study population | Sampling | Risk of bias | Outcome of interest | Summary of findings |
| --- | --- | --- | --- | --- | --- | --- | --- | --- |
| Durski *et al.* [1] | 2017 | Multicentric | Community | General population | Non-probabilistic | Moderate | Case fatality rate | Since 2016, cases have been confirmed in Central African Republic (19 cases), Democratic Republic of the Congo (>1,000 reported per year), Liberia (two), Nigeria (>80), Republic of the Congo (88), and Sierra Leone (one). The reemergence of monkeypox is a global health security concern. |
| Berthet *et al.* [2] | 2010 | NR | Community | General population | Non-probabilistic | Low | Severity and case fatality rates | In June 2010, two teenage boys in the Central African Republic developed pustular skin lesions after eating a wild rodent, later confirmed as monkeypox virus (identical to the DRC strain from a 2001 outbreak). Both recovered after isolation and treatment, highlighting monkeypox’s zoonotic risk in forested regions. |
| Kalthan *et al.* [3] | 2016 | Region 4 | Community | General population | Non-probabilistic | Low | Severity, case fatality, and vaccine uptake rates | A study identified 26 monkeypox cases, with the highest attack rates in children (<10 years) and young adults (21–30 years). The overall attack rate was 5 per 1,000 inhabitants, severe disease occurred predominantly (87.5% of cases) in unvaccinated younger individuals. |
| Kalthan *et al.* [4] | 2015 | Region 6 | Community | General population | Non-probabilistic | Low | Severity and case fatality rates | A 2015–2016 monkeypox outbreak in Bangassou, Central African Republic, affected 12 patients (mostly adults aged 31–40 and children under 10), with a 25% fatality rate (67% in children). The disease, marked by fever, rash, and lymphadenopathy (54.5%), had an attack rate of 0.2/1,000 inhabitants, highlighting the need for isolation, community education, and animal reservoir surveillance to curb transmission. |
| Nakoune *et al.* [5] | 2016 | Region 6 | Community | General population and healthcare workers | Non-probabilistic | Moderate | Severity and case fatality rates | A 2015/2016 familial monkeypox outbreak in Central African Republic infected 10 individuals through household, healthcare, and transport-related transmission. The Zaire genotype strain caused characteristic fever and skin lesions, with two fatal pediatric cases highlighting the disease's severity in children. |
| Besombes *et al.* [6] | 2018 | Region 1 | Community | General population and healthcare workers | Non-probabilistic | Moderate | Severity, case fatality, and vaccine uptake rates | In September 2018, a monkeypox outbreak involving an Aka Pygmy family in Central African Republic began when a 25-year-old woman developed symptoms after butchering wild animals, leading to three waves of intrafamilial transmission that ultimately infected five family members. While PCR confirmed six cases (including three children), serologic evidence suggested prior Orthopoxvirus exposure may have limited secondary spread, highlighting the role of zoonotic exposure and the waning protection from historic smallpox vaccination campaigns. |
| Besombes *et al.* [7] | 2022 | Region 2 | Community | General population | Non-probabilistic | Low | Severity and case fatality rates | A November 2021 monkeypox outbreak in Central African Republic originated from a hunter's contact with a primate, sparking four transmission waves across two families with a 59.5% secondary attack rate (14 confirmed cases). The clade I virus caused severe complications (63.2% of cases) including bronchopneumonia and skin sequelae, with 4% mortality, demonstrating both the high transmissibility and clinical severity of endemic strains that risk international spread |
| ^1^ Date of study completion; CFR: Case fatality rate; NR: Not reported | | | | | | | | |

Supplementary Table 2 Searching Strategies for online databases

| Database | Search Term | Results |
| --- | --- | --- |
| PubMed | (  ("Monkeypox"[Mesh] OR "monkeypox"[tiab] OR "monkeypox virus"[tiab] OR "human monkeypox"[tiab] OR "Mpox"[tiab] OR "mpox"[tiab] OR "MPX"[tiab])  AND  ("Epidemiology"[Mesh] OR "epidemiological"[tiab] OR "Disease Outbreaks"[Mesh] OR "Public Health Surveillance"[Mesh] OR "surveillance"[tiab] OR "Clinical Characteristics"[tiab] OR "Severity of Illness Index"[Mesh] OR "severe"[tiab] OR "Mortality"[Mesh] OR "death"[tiab] OR "Fatality"[tiab] OR "vaccine"[tiab] OR "vaccination"[tiab])  AND  ("Central African Republic"[Mesh] OR "CAR"[tiab])  ) | 15 |
| ScienceDirect | (  ("monkeypox" OR "mpox" OR "MPX")  AND  ("epidemiology" OR "surveillance" OR "characteristic" OR "clinical characteristic" OR "severe" OR "mortality" OR "death" OR “vaccine” OR “vaccination”)  AND  ("CAR" OR "Central African Republic")  ) | 404 |
| Scopus | (TITLE-ABS-KEY(  (monkeypox OR "monkeypox virus" OR "human monkeypox" OR Mpox OR mpox OR MPX)  AND  (“epidemiology” OR “surveillance” OR “characteristics” OR "clinical characteristics" OR “severe” OR “mortality” OR “death” OR “vaccine” OR “vaccination”)  AND  (“CAR” OR “Central African Republic”)  ) | 25 |
| Web of Science | (  ("monkeypox" OR "mpox" OR "MPX")  AND  ("epidemiology" OR "surveillance" OR "characteristic" OR "clinical characteristic" OR "severe" OR "mortality" OR "death" OR “vaccine” OR “vaccination”)  AND  ("CAR" OR "Central African Republic")  ) | 21 |
| Embase | (  ("monkeypox" OR "mpox" OR "MPX")  AND  ("epidemiology" OR "surveillance" OR "characteristic" OR "clinical characteristic" OR "severe" OR "mortality" OR "death" OR “vaccine” OR “vaccination”)  AND  ("CAR" OR "Central African Republic")  ) | 127 |
| Cochrane Library | (  ("monkeypox" OR "mpox" OR "MPX")  AND  ("epidemiology" OR "surveillance" OR "characteristic" OR "clinical characteristic" OR "severe" OR "mortality" OR "death" OR “vaccine” OR “vaccination”)  AND  ("CAR" OR "Central African Republic")  ) | 1 |
| African Journals Online (AJOL) | monkeypox OR mpox OR MPX AND epidemiology OR surveillance OR characteristic OR clinical characteristic OR severe OR mortality OR death OR vaccine OR vaccination AND CAR OR Central African Republic | 2 |
