## Additional Files 2 for "Mpox clinical and epidemiological patterns in the Central African Republic: a systematic review and meta-analysis"

**Period**

**Severity rate (%)**

**Event rate (%)**

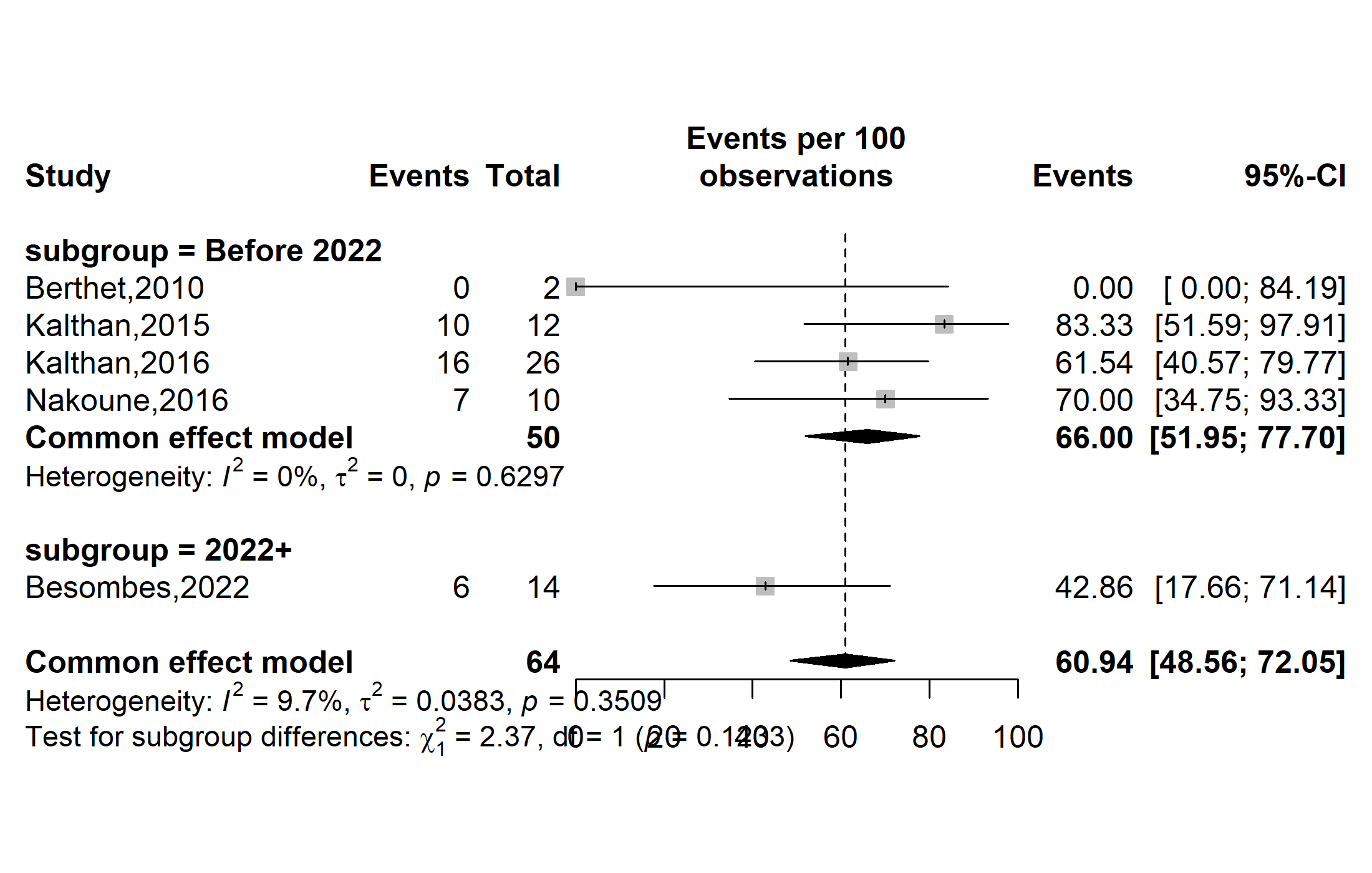

Supplementary Fig. 1 Pooled severity rate by study period among confirmed Mpox cases in Central African Republic (CAR) (*the period represents before and since the 2022 pandemic*)

**Region**

**Event rate (%)**

**Severity rate (%)**

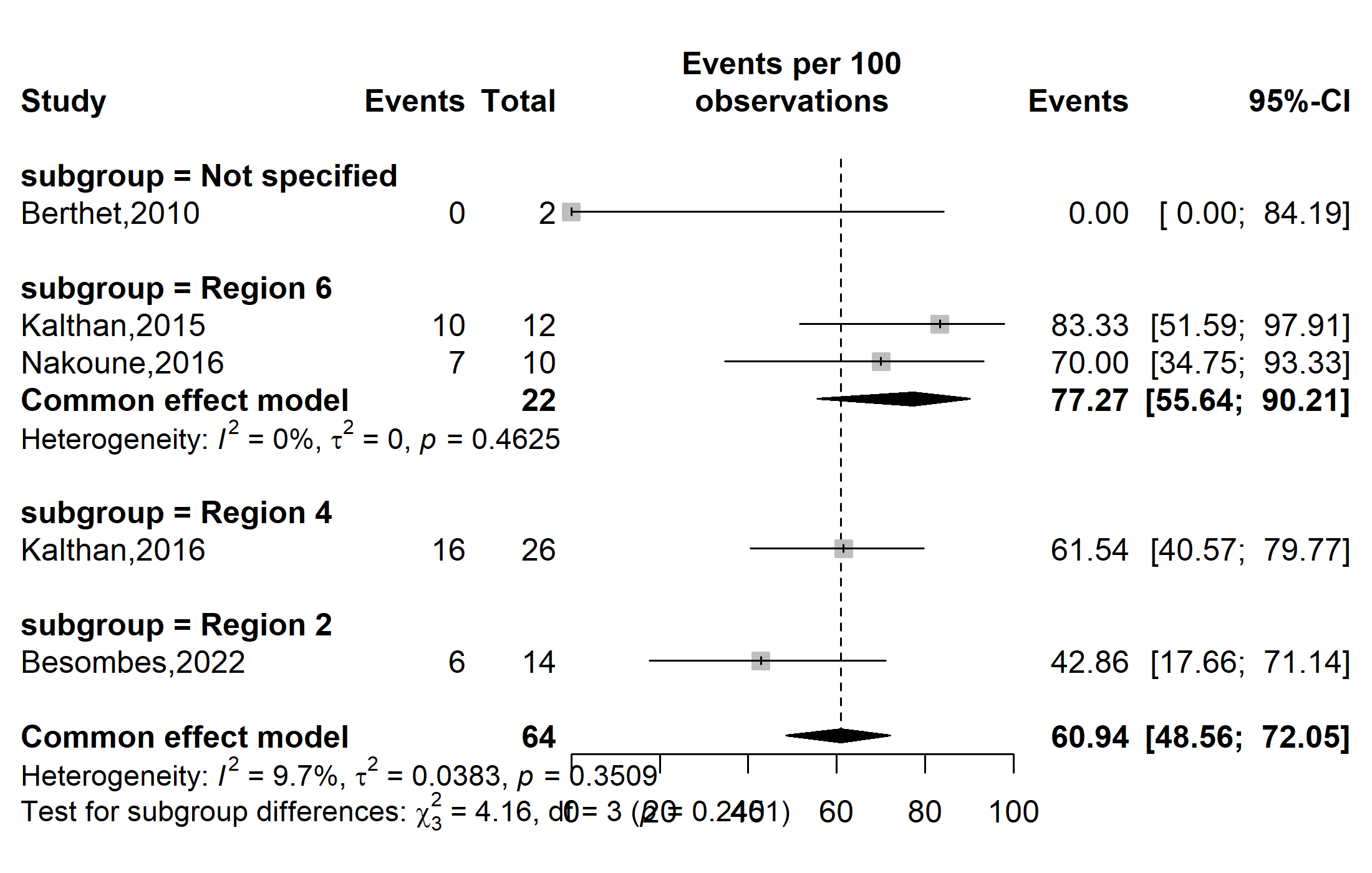

Supplementary Fig. 2 Pooled severity rate by region among confirmed Mpox cases in CAR

**Participant**

**Event rate (%)**

**Severity rate (%)**

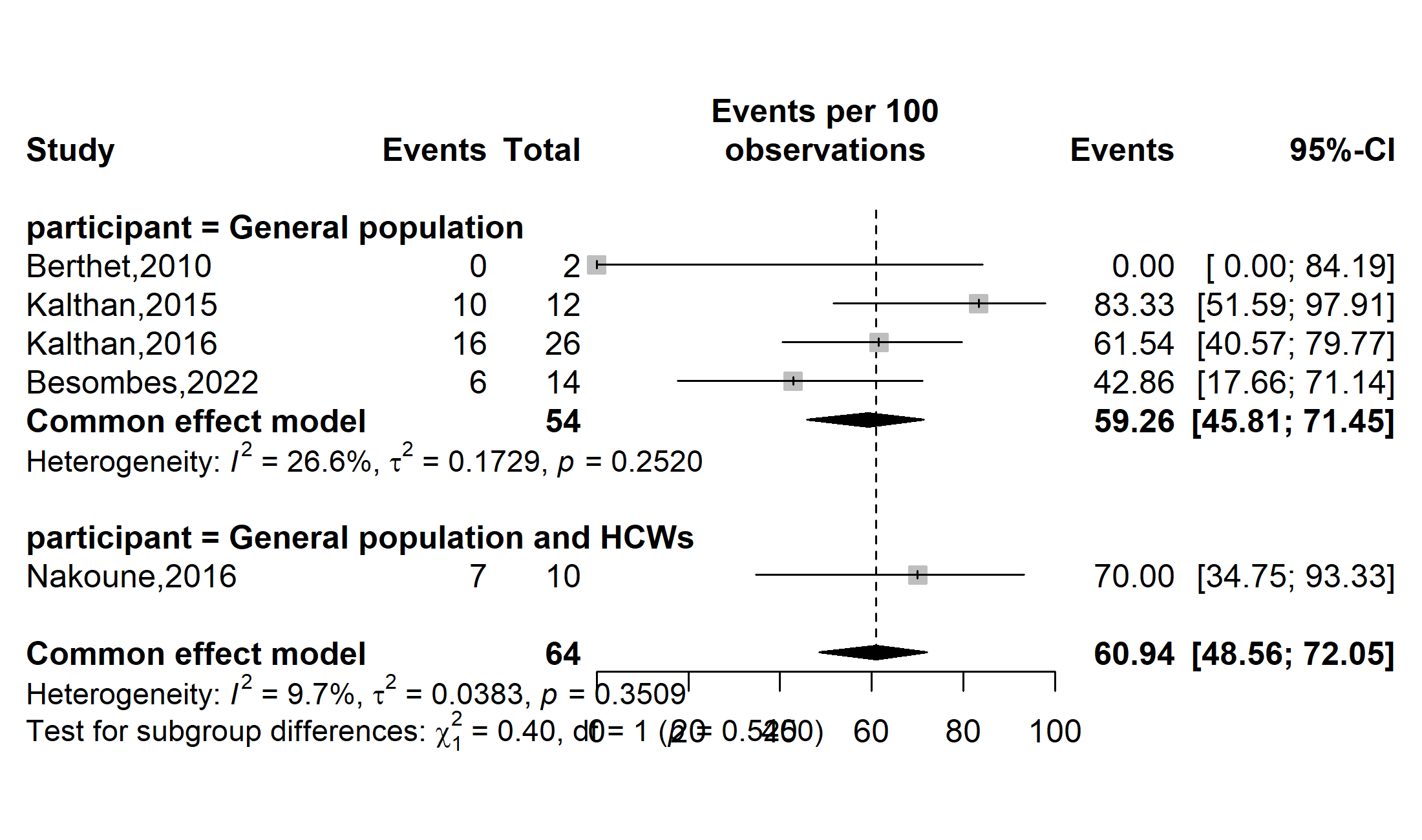

Supplementary Fig. 3 Pooled severity rate by type of participants involved among confirmed Mpox cases in CAR (HCWs: Healthcare workers)

**Publication bias**

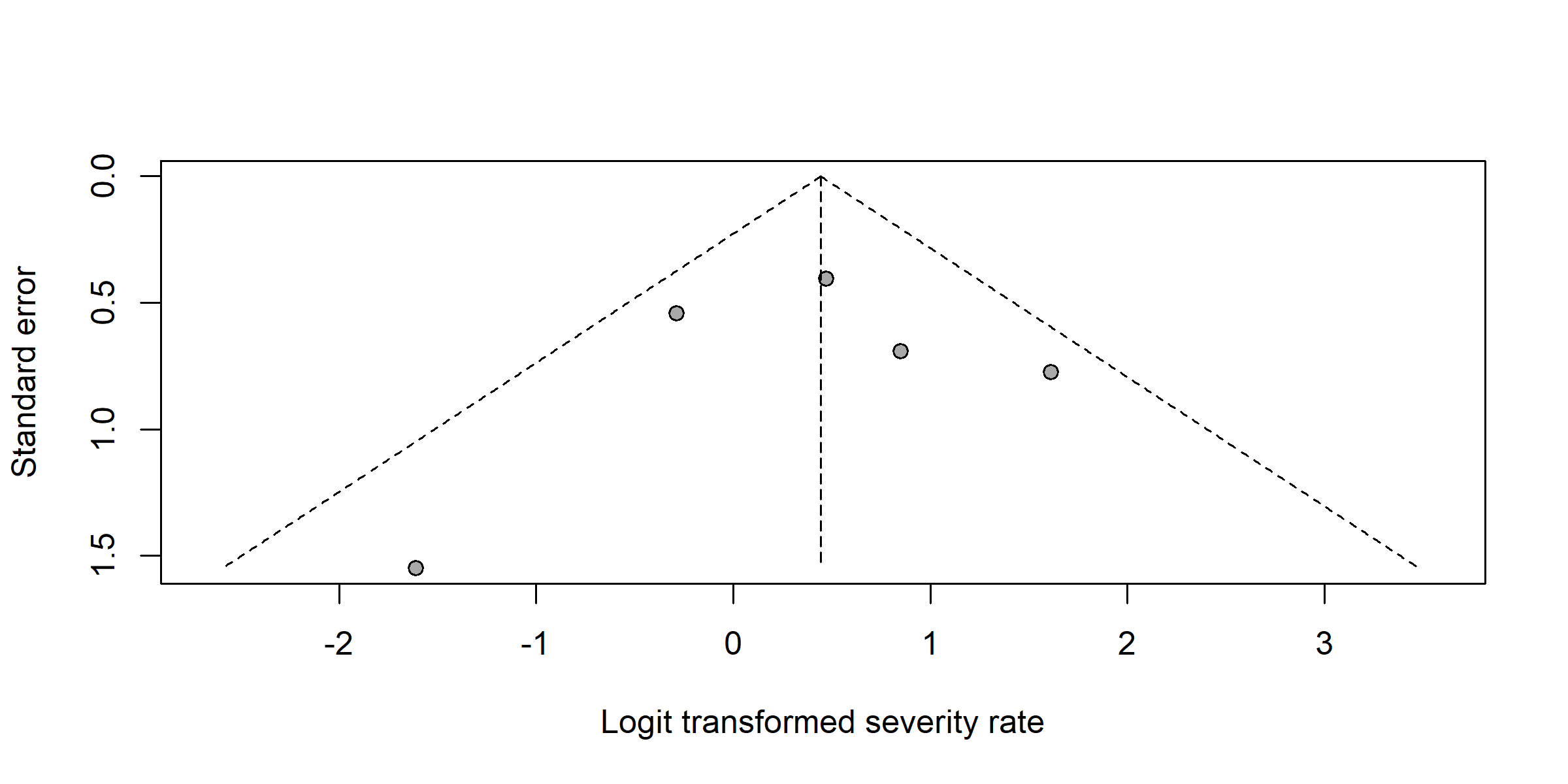

Supplementary Fig. 4 Funnel plot with pseudo 95% confidence limits and tests assessing the publication bias of studies included

**Sensitivity analysis**

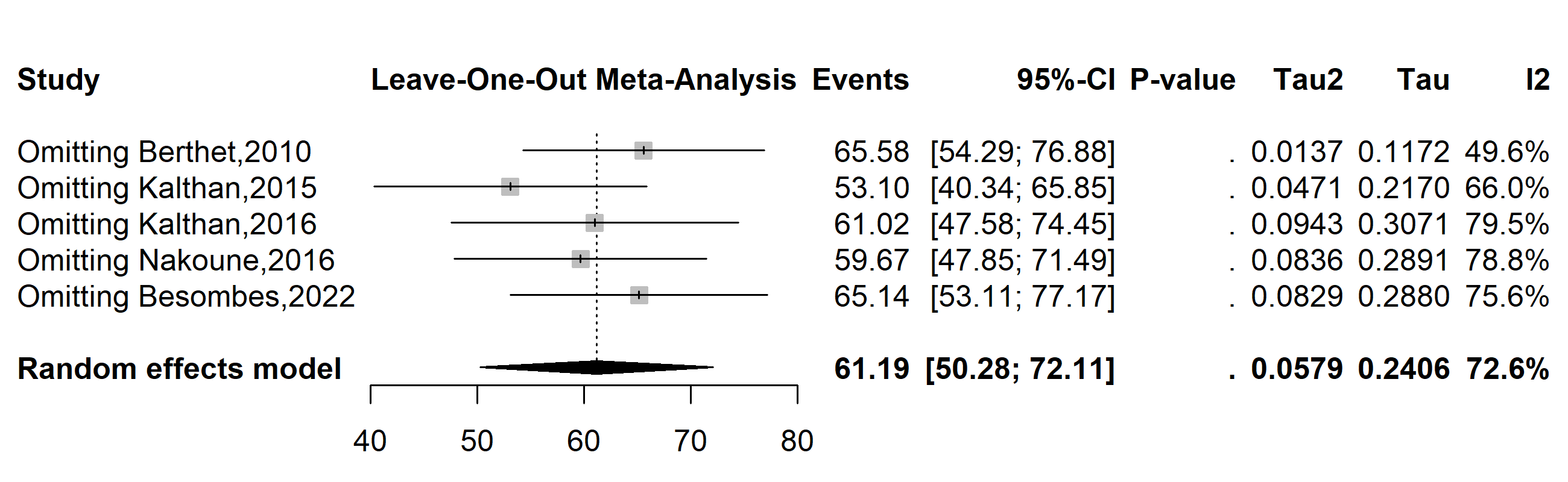

Supplementary Fig. 5 Sensitivity analysis of the Mpox severity rate pooled estimate among confirmed cases in CAR
